# Polygenic risk scores lack prognostic value for adults with severe mental illness

**DOI:** 10.1101/2021.03.19.21253906

**Authors:** Isotta Landi, Deepak Kaji, Liam Cotter, Tielman Van Vleck, Gillian Belbin, Michael Preuss, Ruth Loos, Eimear Kenny, Benjamin S Glicksberg, Noam Beckmann, Paul O’Reilly, Eric E. Schadt, Eric D. Achtyes, Peter F. Buckley, Douglas Lehrer, Dolores P. Malaspina, Steven A. McCarroll, Mark H. Rapaport, Ayman H. Fanous, Michele T. Pato, Carlos N. Pato, Genomic Psychiatry Cohort (GPC) Investigators, Tim B. Bigdeli, Girish N Nadkarni, Alexander W. Charney

## Abstract

Schizophrenia (SCZ) is the archetypal severe mental illness and one of the most deeply characterized human genetic traits. Like most common diseases SCZ is highly polygenic, and as such its genetic liability can be summarized at the individual level by a polygenic risk score (PRS). Polygenic risk scores are a cornerstone of the precision medicine vision, as it is widely anticipated they will come to serve as biomarkers of disease and poor outcomes in real-world clinical practice. However, to date, few studies have assessed their actual prognostic value relative to current standards-of-care. SCZ is an ideal test case towards this end because the predictive power of the SCZ PRS exceeds that of most other common diseases. Here, we analyzed clinical and genetic data from two multi-ethnic cohorts totaling 8,541 adults with SCZ and related psychotic disorders, assessing whether the SCZ PRS improves poor outcome prediction relative to clinical features captured in a standard psychiatric interview. For all outcomes investigated, the SCZ PRS did not improve the performance of predictive models, an observation that was generally robust to divergent case definitions and ancestral backgrounds of study participants. These findings demonstrate the limited potential of even the most powerful contemporary polygenic risk scores as a tool for individualized outcome prediction.

---

Schizophrenia (SCZ) is the archetypal severe mental illness and one of the most deeply characterized human genetic traits. Like most common diseases SCZ is highly polygenic, and as such its genetic liability can be summarized at the individual level by a polygenic risk score (PRS)^1,2^. Polygenic risk scores are a cornerstone of the precision medicine vision, as it is widely anticipated they will come to serve as biomarkers of disease and poor outcomes in real-world clinical practice^3–5^. However, to date, few studies have assessed their actual prognostic value relative to current standards-of-care^6^. SCZ is an ideal test case towards this end because the predictive power of the SCZ PRS exceeds that of most other common diseases^6–17^. Here, we analyzed clinical and genetic data from two multi-ethnic cohorts totaling 8,541 adults with SCZ and related psychotic disorders, assessing whether the SCZ PRS improves poor outcome prediction relative to clinical features captured in a standard psychiatric interview. For all outcomes investigated, the SCZ PRS did not improve the performance of predictive models, an observation that was generally robust to divergent case definitions and ancestral backgrounds of study participants. These findings demonstrate the limited potential of even the most powerful contemporary polygenic risk scores as a tool for individualized outcome prediction.

In real-world clinical practice, there are few if any objective metrics used to predict outcomes for adults with mental illness^18^. Instead, for a given patient, treating clinicians gather information from routine psychiatric interviews, forming a gestalt of the clinical trajectory that then informs clinical decision-making. To investigate the prognostic utility of the SCZ PRS compared to this standard-of-care, the first challenge was to represent this standard-of-care in an analytic framework. Towards this end, we integrated clinical and genetic data from Bio*Me*, an ancestrally diverse biobank for translational research in a large New York City health system (***Table 1*** and ***Supplementary Table 5***)^19^. Case definitions, outcomes, and clinical features potentially predictive of outcomes were sourced from structured electronic medical record (EMR) data, rule-based natural language processing of unstructured clinical notes, and a biobank survey of general health (***Supplementary Table 1***). Cases were defined as any Bio*Me* participant who had been billed for treatment of SCZ or a related psychotic disorder on at least one occasion (N=762). This broad definition was intended to capture the spectrum of psychosis encountered in real-world clinical settings (***Supplementary Table 4***). SCZ polygenic risk scores, calculated using summary statistics from a large SCZ genome-wide association study (GWAS)^14^, explained a significant proportion of variance in case status (Nagelkerke’s pseudo-R^2^ = 0.006, p-value = 1.49 × 10^−9^, ***Supplementary Table 3***). A set of clinical features was defined to encompass the symptoms, comorbidities, and other biopsychosocial data elements that might be predictive of poor outcomes in adults with psychotic illness (***Supplementary Table 1)***.

We initially defined a set of six outcomes that could be captured in the data sources available as proxies for a poor clinical course: (1) requiring inpatient psychiatric treatment, (2) prescription of two or more unique antipsychotics, (3) prescription of clozapine, (4) aggressive behavior, (5) self-injurious behavior, and (6) homelessness. The variance explained by the SCZ PRS was assessed for each of these outcomes in Bio*Me*, and after adjusting for multiple tests a significant association was observed for inpatient psychiatric treatment (Nagelkerke’s pseudo-R^2^=0.015, adjusted p-value = 0.02) and aggressive behavior (Nagelkerke’s pseudo-R^2^=0.026, adjusted p-value = 0.02) (***Supplementary Table 3***). Since, by definition, the SCZ PRS would only have predictive ability for statistically associated outcomes, subsequent predictive modeling analyses were limited to these two outcomes.

To predict these two outcomes, Bio*Me* cases were split into training (i.e., discovery) and test (i.e., replication) sets, which included 70% and 30% of cases, respectively (***Supplementary Table 6***). Three configurations of a logistic regression model were then fit to the training data and compared to one another. Each configuration used a different set of features to predict the outcome variable: clinical features only, clinical features plus SCZ PRS, and SCZ PRS only. The two configurations that included the SCZ PRS also included the top 20 genetic principal components (PCs) to account for the confounding effects of ancestry. For each configuration, performance was initially assessed in the training data alone, using a repeated cross-validation framework wherein the training data was iteratively split into one part for model fitting and another part for performance evaluation. *F*_2_ scores were used as the metric of evaluation for this initial performance assessment. *F*_2_ scores combine precision (i.e., a metric used to limit the number of false positives) and recall (i.e., a metric used to limit the number of false negatives), but with greater weight on recall. Compared to other evaluation metrics, when dealing with imbalanced classes, as was the case here, *F*_2_ scores prevent a high false negative rate from creating the appearance of good performance. For the two outcomes considered in Bio*Me*, model performance did not differ in the training step between the configuration that used clinical features alone and the configuration that used clinical features plus the SCZ PRS (***Table 2 and Figure 1a,c***). Both of these configurations outperformed the configuration with the SCZ PRS alone (*p*_*s*_ < 0.001; ***Supplementary Table 8***).

To assess the generalizability of the three model configurations, each was then fit to the entire training set and evaluated on the test set. In addition to the *F*_2_ score, to assess generalizability we also used the area under precision-recall curve (AUPRC). The latter allowed us to assess how performance changes as the level of confidence required to predict the outcome is varied. AUPRC estimates for the three configurations were compared to one another via bootstrap resampling of the test set. No difference in generalizability was observed between the configuration with clinical features only and the configuration with clinical features plus the SCZ PRS for either aggressive behavior (*F*_2_ = 0.44; *AUPRC* (*sd*) = 0.71 (0.08) for both configurations) or inpatient psychiatric admission (*F*_2_ = 0.67; *AUPRC* (*sd*) = 0.77 (0.05) for clinical features only; *F*_2_ = 0.68; *AUPRC* (*sd*) = 0.73 (0.06) for clinical features plus SCZ PRS; ***Figure 1b,d***; ***Supplementary Table 8***).

The results of both the testing and training performance assessments were generally consistent across the three main ancestry groups (i.e., African [AFR], admixed American [AMR], and European [EUR]) in Bio*Me* (***Table 2*** and ***Supplementary Figures 3-5***). The only exception was aggressive behavior in cases of EUR ancestry. The number of EUR subjects that developed aggressive behavior was very low for both training and test sets (*N* = 5 (7*%*) and *N* = *2* (6*%*), respectively; ***Supplementary Table 7***), and this likely explains the poor performance during training, very high standard deviations, and low or null generalization scores regardless of the configuration (***Table 2*** and ***Supplementary Figure 5***). From earlier work, it is known that SCZ polygenic risk scores in the top decile have greater predictive capacity in case-control comparisons^20^. We therefore repeated the predictive modeling analyses using a binarized version of the SCZ PRS instead of the continuous variable, where a value of one indicated the SCZ PRS fell within the top decile in Bio*Me* and a value of zero indicated it did not. This did not appreciably change any of the observations (***Supplementary Table 8 and Supplementary Figure 1c***,***h***,***e***,***j***).

To qualitatively replicate the prognostic performance of the SCZ PRS seen in Bio*Me*, we next integrated clinical and genetic data from 7,779 cases in the multiethnic Genomic Psychiatry Cohort (GPC)^21^. In contrast to Bio*Me*, diagnoses, clinical features, and outcomes for the GPC were established using the Diagnostic Interview for Psychosis and Affective Disorders (DIPAD), a validated instrument based on the OPCRIT system^22^ (***Table 1*** and ***Supplementary Table 5***). Details of case ascertainment, DIPAD assessments, genotyping, and polygenic risk scoring for GPC have been previously described^23^. For the present study, five DIPAD items were treated as outcomes: “Suicidality” (OPCRIT item 43), “Impairment or incapacity during disorder” (OPCRIT item 87), “Deterioration from premorbid level of functioning” (OPCRIT item 88), “Psychotic symptoms respond to antipsychotic medications” (OPCRIT item 89), and “Course of disorder” (OPCRIT item 90). OPCRIT items 87-89 were excluded due to little variation in our cohort. That is, nearly all GPC cases were incapacitated while ill, deteriorated from premorbid level of functioning, and had some response to treatment. Of the remaining two outcomes, the SCZ PRS was observed to explain a significant proportion of variance only for the course of disorder (Nagelkerke’s pseudo-R^2^=0.002, adjusted p-value = 3.49 × 10^−3^; ***Supplementary Table 3***), which had been binarized because nearly all GPC cases fell into the two most severe categories: “continuous chronic illness” and “continuous chronic illness with deterioration”.

The same three-configuration predictive modeling framework described for Bio*Me* was used to predict the course of disorder in GPC. Clinical predictors included all items in the DIPAD besides the outcome (***Supplementary Table 2***). In the training step, a statistically significant difference in model performance was observed between the configuration that used clinical features alone and the configuration that used clinical features plus the SCZ PRS (training estimates *F*_2_ (*sd*) = 0.555 (0.015) for clinical features only; *F*_2_ (*sd*) = 0.560 (0.015) for clinical features plus SCZ PRS). This small difference, though statistically significant, was unlikely due to an actual contribution of the PRS to outcome prediction. The logistic regression models used during the training step tended to underfit with less regularization, suggesting they were too simple and that performance was improved simply by adding complexity through additional features (***Supplementary Figure 1k-o*** and ***Table 2***). This conclusion was further supported by the fact that no difference in generalizability was observed between the configuration with clinical features only and the configuration with clinical features plus the SCZ PRS (*F*_2_ = 0.56; *AUPRC* (*sd*) = 0.65 (0.01) for clinical features only; *F*_2_ = 0.57; *AUPRC* (*sd*) = 0.66 (0.01) for clinical features plus SCZ PRS; *p_s_* = 0.60; ***Table 2 and Supplementary Table 8***). Stratifying GPC cases by ancestry generally showed results consistent with the analysis of the full cohort. There were no significant differences observed in the generalizability of the clinical only and clinical plus SCZ PRS model configurations for any ancestral subgroup (***Table 2***). Binarizing the SCZ PRS based on the top decile in GPC did not improve the generalizability of configurations that included the SCZ PRS compared to clinical features only in the full, AFR ancestry, or EUR ancestry cohorts. For the AMR ancestry cohort, binarization yielded a statistically significant difference in the generalizability of the configuration with clinical features plus SCZ PRS compared to the clinical features only configuration (*AUPRC* (*sd*) = 0.6*2*3 (0.045); *AUPRC* (*sd*) = 0.655 (0.045); *p* < 0.0001; ***Supplementary Table 8***), likely due to the reduced sample size relative to the other ancestry groups in GPC (***Supplementary Table 7***).

There are several limitations to the approach employed here to assess the ability of polygenic risk scores to predict poor outcomes in adults with severe mental illness. First, PRS analyses can be highly confounded by the ancestral backgrounds of the discovery and target cohorts. In this report, the latter were ancestrally diverse while the former was predominantly of EUR ancestry^23,24^. The steps we took computationally to account for the potential confounding effects of ancestry appear to have been sufficient, as the model configurations that included the SCZ PRS did not perform better in the EUR subsets of Bio*Me* and GPC compared to other ancestries. Second, the summary statistics used to calculate the SCZ PRS in Bio*Me* and GPC were derived from GWAS that compared SCZ cases to controls, as adequately powered GWAS of outcomes amongst SCZ cases are lacking. To mitigate this limitation, we only performed predictive modeling on outcomes that had a statistically significant association with the SCZ PRS. Third, our study design did not afford us the opportunity to account for the temporal relationship between clinical predictors and outcomes. As the symptomatology of psychosis is highly stable over time^25^, we do not expect the inability to temporally link clinical predictors (which were mostly symptoms) to outcomes confounded the primary conclusions of the study. Fourth, our replication analysis was qualitative in nature. Bio*Me* and GPC case cohorts were ascertained using divergent strategies, resulting in different sets of clinical variables for analysis. Bio*Me* cases are representative of patients treated for psychosis in real-world clinical settings, and GPC cases of a more rigidly defined SCZ clinical research cohort. On the one hand, this prevented us from performing a strict replication analysis. On the other hand, analyses of outcomes in both cohorts concluded that the SCZ PRS did not improve prediction, suggesting the result is robust.

Polygenic risk scores are widely viewed as a means to realize one of the central features of the precision medicine vision, namely the routine use of genetics to personalize care^3–5^. This view has taken hold despite statistical geneticists having long urged caution given the low variance in phenotype explained by the PRS^6^. We suspect the lack of prognostic value reported here for the SCZ PRS, therefore, may come as a surprise to some but not others. Cross-talk between clinical and genetic experts will be critical moving forward in order establish realistic expectations regarding the extent to which genetics will reshape healthcare. While this study was limited to SCZ and related psychotic disorders, the implications extend across medicine broadly since the variance in phenotype explained by the SCZ PRS exceeds that of most other diseases^7–17^. Indeed, our findings extend to mental illness similar observations made recently in cardiovascular disease^26^, and further show that the poor prognostic performance of the PRS is not a function of the ancestral background of the study population.

Future studies on the clinical utility of polygenic risk scores in the care of the mentally ill should focus on predicting disease onset amongst individuals at high risk rather than predicting outcomes amongst those with established illness. Earlier studies towards this end have shown promise^27^. Performing these investigations at the scale needed to establish clinical utility will likely require a reprioritization of current research strategies, emphasizing deep phenotyping of individuals that have already been genotyped over additional genotyping of poorly phenotyped cohorts. Though we did not find evidence to support a clinical role for the PRS, we did find untapped translational potential for clinical data analytics. Our analyses in both BioMe and GPC suggest that information obtained during the course of routine psychiatric care has value for predicting outcomes. In parallel to efforts for biomarker discovery, therefore, the field should prioritize the standardization of clinical data collection and storage at the point-of-care. Doing so will enable the development of predictive models that can be built into EMR systems to optimize care pathways for SCZ and other mental illnesses.

**Table 1.** Bio*Me* and Genomic Psychiatry Cohort (GPC) cohort descriptions. Class counts are reported for investigated outcomes, i.e., development of aggressive behavior (Aggressive), number of psychiatric admissions, and course of disorder.

|  | **BioMe** | **GPC** |
| --- | --- | --- |
| N | 762 | 7,779 |
| Gender (F/M) | 391/371 | 2,592/5185 |
| Ancestry (AFR/AMR/EUR) | 349/316/97 | 3,522/618/3639 |
| Average age yrs (sd) | 59.39 (15.84) | 43.93 (12.51) |
| **Outcome (0/1)** |  |  |
| Aggressive (present/absent) | 681/81 | - |
| Psychiatric admissions<br>(0 admissions/>=1 admissions) | 547/215 | - |
| Course of disorder (w/<br>deterioration; w/o deterioration) | - | 4,307/3,472 |

GPC: Genomic Psychiatry Cohort; F: female; M: male; N: counts; AFR: African ancestry; AMR: admixed American; EUR: European.

**Table 2.**
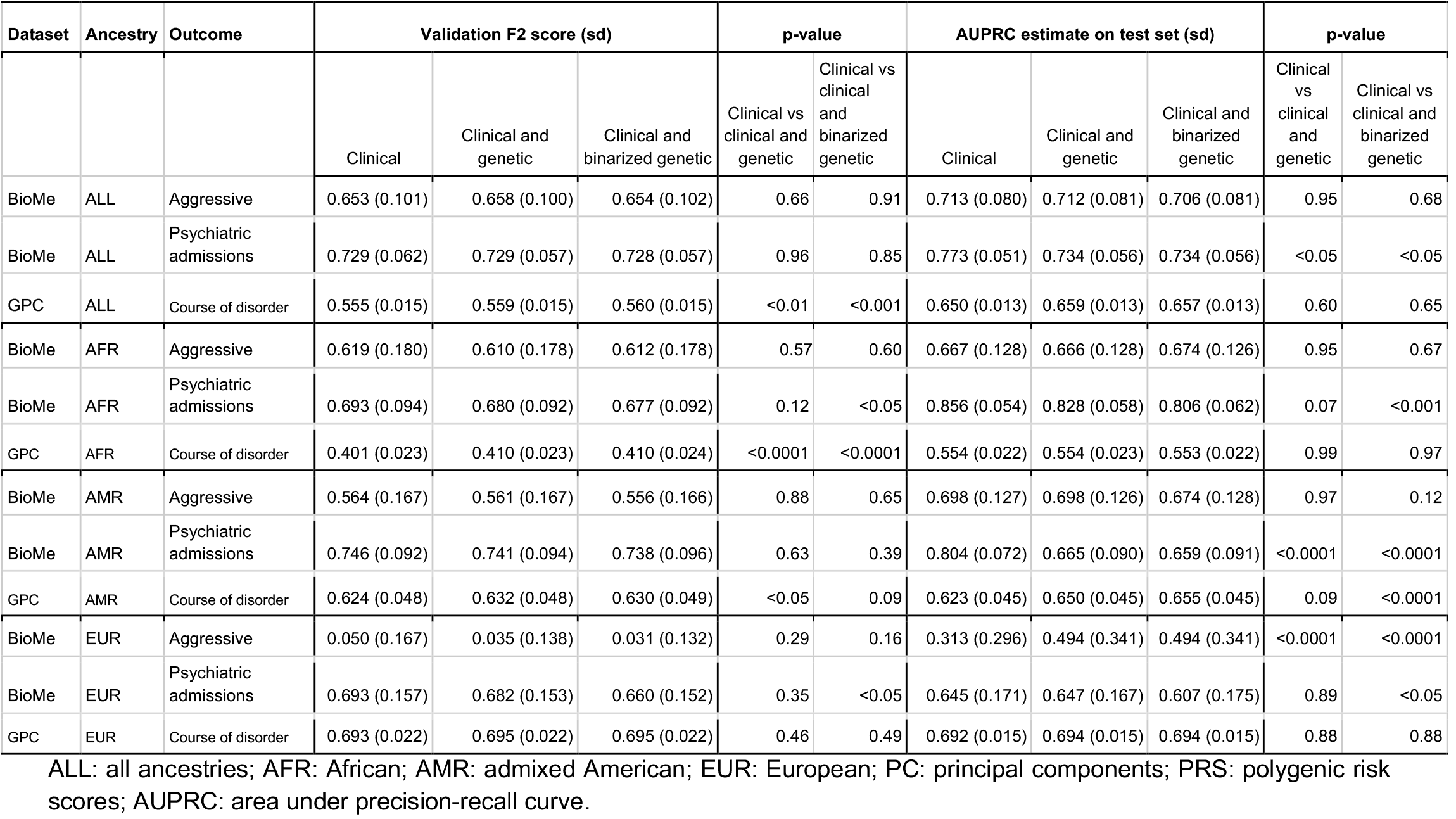
Model prediction performance for models with clinical, clinical and genetic, and clinical and binarized genetic features. *F*_2_ validation and bootstrap estimates of area under the precision-recall curve (AUPRC) on test sets are displayed to investigate models’ performance. P-values refer to two-sided pairwise t-tests with Benjamini-Hochberg correction for significant one-way re peated-measure ANOVAs with different sets of features as groups.

| Dataset | Ancestry | Outcome | Validation F2 score (sd) |  |  | p-value |  | AUPRC estimate on test set (sd) |  |  | p-value |  |
| --- | --- | --- | --- | --- | --- | --- | --- | --- | --- | --- | --- | --- |
|  |  |  | Clinical | Clinical and genetic | Clinical and binarized genetic | Clinical vs clinical and genetic | Clinical vs clinical and binarized genetic | Clinical | Clinical and genetic | Clinical and binarized genetic | Clinical vs clinical and genetic | Clinical vs clinical and binarized genetic |
| BioMe | ALL | Aggressive | 0.653 (0.101) | 0.658 (0.100) | 0.654 (0.102) | 0.66 | 0.91 | 0.713 (0.080) | 0.712 (0.081) | 0.706 (0.081) | 0.95 | 0.68 |
| BioMe | ALL | Psychiatric admissions | 0.729 (0.062) | 0.729 (0.057) | 0.728 (0.057) | 0.96 | 0.85 | 0.773 (0.051) | 0.734 (0.056) | 0.734 (0.056) | <0.05 | <0.05 |
| GPC | ALL | Course of disorder | 0.555 (0.015) | 0.559 (0.015) | 0.560 (0.015) | <0.01 | <0.001 | 0.650 (0.013) | 0.659 (0.013) | 0.657 (0.013) | 0.60 | 0.65 |
| BioMe | AFR | Aggressive | 0.619 (0.180) | 0.610 (0.178) | 0.612 (0.178) | 0.57 | 0.60 | 0.667 (0.128) | 0.666 (0.128) | 0.674 (0.126) | 0.95 | 0.67 |
| BioMe | AFR | Psychiatric admissions | 0.693 (0.094) | 0.680 (0.092) | 0.677 (0.092) | 0.12 | <0.05 | 0.856 (0.054) | 0.828 (0.058) | 0.806 (0.062) | 0.07 | <0.001 |
| GPC | AFR | Course of disorder | 0.401 (0.023) | 0.410 (0.023) | 0.410 (0.024) | <0.0001 | <0.0001 | 0.554 (0.022) | 0.554 (0.023) | 0.553 (0.022) | 0.99 | 0.97 |
| BioMe | AMR | Aggressive | 0.564 (0.167) | 0.561 (0.167) | 0.556 (0.166) | 0.88 | 0.65 | 0.698 (0.127) | 0.698 (0.126) | 0.674 (0.128) | 0.97 | 0.12 |
| BioMe | AMR | Psychiatric admissions | 0.746 (0.092) | 0.741 (0.094) | 0.738 (0.096) | 0.63 | 0.39 | 0.804 (0.072) | 0.665 (0.090) | 0.659 (0.091) | <0.0001 | <0.0001 |
| GPC | AMR | Course of disorder | 0.624 (0.048) | 0.632 (0.048) | 0.630 (0.049) | <0.05 | 0.09 | 0.623 (0.045) | 0.650 (0.045) | 0.655 (0.045) | 0.09 | <0.0001 |
| BioMe | EUR | Aggressive | 0.050 (0.167) | 0.035 (0.138) | 0.031 (0.132) | 0.29 | 0.16 | 0.313 (0.296) | 0.494 (0.341) | 0.494 (0.341) | <0.0001 | <0.0001 |
| BioMe | EUR | Psychiatric admissions | 0.693 (0.157) | 0.682 (0.153) | 0.660 (0.152) | 0.35 | <0.05 | 0.645 (0.171) | 0.647 (0.167) | 0.607 (0.175) | 0.89 | <0.05 |
| GPC | EUR | Course of disorder | 0.693 (0.022) | 0.695 (0.022) | 0.695 (0.022) | 0.46 | 0.49 | 0.692 (0.015) | 0.694 (0.015) | 0.694 (0.015) | 0.88 | 0.88 |

ALL: all ancestries; AFR: African; AMR: admixed American; EUR: European; PC: principal components; PRS: polygenic risk scores; AUPRC: area under precision-recall curve.

**Figure 1.**
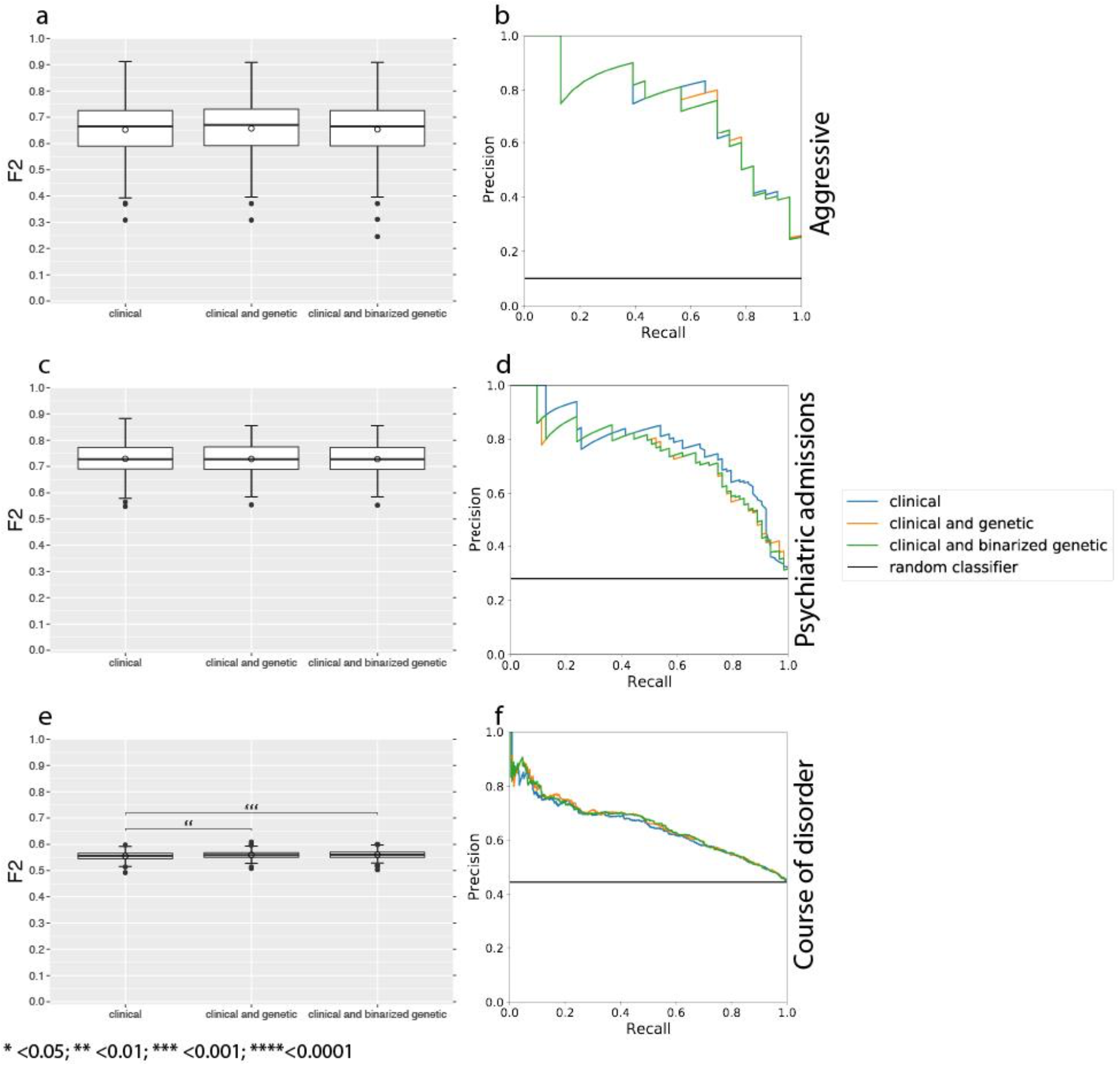
Models’ performance for Bio*Me* datasets and Genomic Psychiatry Cohort (GPC). Average *F*_2_ scores obtained with different feature configurations (i.e., clinical, clinical and genetic, clinical and binarized genetic) in predicting outcomes are displayed in panels (a), (c), and (e). P-value thresholds from two-sided pairwise t-test comparisons with Benjamini-Hochberg correction are displayed for significant results. Panels (b), (d), and (f) display precision-recall curves for linear regression models in predicting outcomes on test sets. Performance of random classifiers is displayed as reference.

## Online Methods

### BioMe - defining cases, outcomes, and clinical variables

Founded in September 2007, Bio*Me* is a biobank that links genetic and electronic medical record (EMR) data for over 30,000 individuals recruited primarily in ambulatory care settings in the Mount Sinai Health System (MSHS) in New York City. The current study was approved by the Icahn School of Medicine at Mount Sinai’s Institutional Review Board (Institutional Review Board 07-0529). All study participants provided written informed consent.

Bio*Me* cases, outcomes, and predictive clinical variables were derived from the EMR and a medical questionnaire that participants complete upon study intake (***Supplementary Table 1)***. Cases were defined by the presence of at least one billing code in the “Schizophrenia, Schizotypal, Delusional, And Other Non-Mood Psychotic Disorders” sub-chapter of the 10th version of the International Classification of Disease (ICD-10)^28^.

Outcomes included proxies for treatment response (the number of psychiatric admissions, the number of unique antipsychotic medications prescribed, a history of being prescribed clozapine), homelessness, and development of aggressive behavior.

Drug prescription data was obtained from the Mount Sinai Data Warehouse (MSDW) as non-standardized character strings (e.g., “CLOZAPINE 12.5 MG DISINTEGRATING TABLET”, “CLOZAPINE 100 MG TAB”). To standardize drug names, we took the original drug name strings from MSDW and ran them through an open-source clinical natural language processing pipeline called CLAMP^29^. The output from CLAMP includes a conversion of identified drug strings to RxNorm Concept Unique Identifiers (RXCUIs). The RXCUI codes from CLAMP required further standardization since they did not all refer to a base ingredient. To further standardize, the RXCUI codes from CLAMP were mapped to the base ingredient information using the RxNorm API. While RxNorm effectively standardizes drug names, such that the two clozapine examples above would both be identified as RXCUI 2626, it does not group drugs into classes. To do this, we mapped RXCUIs to the Anatomical Therapeutic Chemical (ATC) classification system, and any RXCUI mapping to the ATC level 3 code N05A (“Antipsychotics”) was considered an antipsychotic for the purposes of this report. The number of unique antipsychotics was simply defined as the number of unique drugs in this class an individual had been prescribed on at least one occasion. For example, if an individual had been prescribed clozapine on 12 separate occasions and aripiprazole on three separate occasions, the total number of unique antipsychotics prescribed for this individual would be two. We used a threshold of two unique antipsychotics to binarize this variable, as we have shown previously that this approach is a valid proxy for poor treatment response in observational drug registry studies^30^.

Predictive clinical variables were selected to be representative of information typically ascertained during a routine psychiatric history and mental status exam, and included symptoms (e.g., hallucinations, delusions), family history, comorbidities, and risk factors previously established through epidemiological studies of SCZ (e.g., migration status, substance use).

Case status and a subset of the outcomes and predictive clinical variables could be extracted from structured tabular data (either from the EMR or the Bio*Me* questionnaire) but as most of this information exists only in the form of unstructured clinical notes it was necessary to use natural language processing (NLP) to extract the information. The CLiX NLP engine utilizes description logic from the Systematized Nomenclature of Medicine Clinical Terms (SNOMED-CT), a compendium of hundreds of thousands of medical concepts organized in a hierarchical fashion with respect to one another. For the current report, we manually curated a set of over 100 concepts relevant to psychotic illness from the prompts of rating scales commonly used in psychosis research, covering both symptoms (e.g., hallucinations) as well as important biopsychosocial features (e.g., “living on the street”). These terms were then manually mapped to the corresponding SNOMED-CT code, and we assessed the ability of CLiX to accurately detect the presence of these SNOMED-CT concepts in Bio*Me* clinical notes. For each concept, we randomly selected for manual review by a clinical psychiatrist (AWC) five patient records with a positive query. A positive query was said to be a true positive if the symptom was being described by the author of the note as present in the patient, and otherwise was defined as a false positive. Terms with a positive predictive value (PPV) below 80% after review of five records were discarded. For the remaining 60 terms an additional 20 records were manually reviewed, for a total of 1,790 clinical notes manually reviewed for this report. Terms with a PPV over 90% after review of 25 notes were retained for downstream analysis. After collapsing terms representing the same concept (e.g., “fast flow of thought” and “racing thoughts”) a set of 32 high-confidence variables derived from NLP were retained.

### BioMe - genotyping and quality control

Genotyping and quality control (QC) for Bio*Me* participants has been previously described^31^. In brief, Bio*Me* participants (N=32,595) were genotyped on the Illumina Global Screening Array (GSA) platform (635,623 variants). All QC steps were conducted using PLINK (v1.90b3.43)^32,33^. QC was performed stratified by self-reported race/ethnicity categories. Individuals were filtered according to heterozygosity rate (removed if +/6 standard deviations of the population-specific mean), call rate (<95%), discordant self-reported and genetically-determined sex, and duplicates. Sites were filtered for call rate (below 95% were excluded), and violation of Hardy-Weinberg equilibrium (HWE) (threshold of p <1×10^−5^ for all populations except HL, where it was set to p < 1×10^−13^). After these QC steps, 31,705 individuals and 604,869 sites remained for downstream analysis. To estimate genetic relatedness, pairwise kinship coefficients were then estimated for all remaining participants using all SNPs using the KING software (v1.4)^34^ by passing the --kinship flag. For any first or second-degree relationships (as defined by a pairwise kinship coefficient of >= 0.0442 in the KING output), we randomly removed one individual from the analysis (leading to the exclusion of 3,500 participants).

### BioMe - phasing and imputation

Prior to phasing, additional QC steps were applied. SNP positions were lifted over to GRCh37/hg19, SNPs were removed for call rate below 99%, MAF <1% (n=135,011), and being palindromic. The data was subsequently merged with the Thousand Genomes Project (TGP) reference panel (N=2,504 individuals)^35^, and only intersecting sites were retained. Phasing was performed using SHAPEIT (v2.r790)^36^ using the HapMap2 genetic map (build: GRCh37/hg19), default parameters, and –output-max –force. Imputation was performed on phased haplotypes using IMPUTE(v2.3.2)^37^ with the TGP phase III data release as the reference panel, and the addition of the following flag: “-filt_rules_1 ‘ALL<0.0002’ ‘ALL>0.9998’.

### BioMe - principal component analysis

Principal components analysis (PCA) was performed in Bio*Me* using PLINK (v1.90b6.10) in order to account for ancestry in downstream analyses^38^. Prior to PCA we restricted analysis to autosomal sites with a minor allele frequency (MAF) in Bio*Me* greater than 0.05. We also removed regions of the genome known to be under recent selection, specifically *HLA* (chr6:27000000-35000000, hg19), *LCT* (chr2:135000000-137000000), an inversion on chromosome 8 (chr8:6000000-16000000), a region of extended LD on chromosome 17 (chr17:40000000-45000000), *EDAR* (chr2:109000000-110000000), *SLC2A5* (chr15:48000000-49000000), and *TRBV9* (chr7:142239536-142240057). Pruning was then performed using the ‘--indep-pairwise’ option in PLINK with a window size of 1000 kilobases and a pairwise r^2^ threshold of 0.1, resulting in 76,992 sites that were then used to calculate the first 20 principal components (PCs).

### BioMe - genetic ancestry determination

We utilized genetic ancestry designations generated in a previous Bio*Me* report^31^. In brief, pairwise genomic tracts inherited identity-by-descent (IBD) were identified between Bio*Me* participants and 2,504 TGP participants. This information was used to construct a network of pairwise IBD sharing between all individuals who were not directly related. To identify ‘communities’ of individuals enriched for shared genealogical ancestry community detection via flow-based clustering using the InfoMap algorithm^39,40^. For the current report, we only included Bio*Me* individuals who fell into one of 17 distinct IBD communities containing at least 100 individuals. As previously reported, these communities represent geographical or ethnic substructure within the Bio*Me* population, labels that were assigned systematically using self-reported information about geographical origins or ethnicity. For the ancestry-stratified analyses in this report, the geographic IBD community labels were further collapsed into EUR, AMR and AFR groupings manually.

### BioMe - polygenic risk scoring

Polygenic risk scores for SCZ were calculated for each of the individuals in Bio*Me* with a self-reported ancestry data point. As a discovery dataset we used the Psychiatric Genomics Consortium Wave 3 schizophrenia (PGC3SCZ) genome-wide association study meta-analysis summary statistics (v2). To account for the known confounding effects of ancestry on PRS calculations, a multi-step procedure was followed to calculate scores separately for each ancestry in Bio*Me* (with ancestry assigned as described above). First, we identified the set of SNPs present in all three of the imputed Bio*Me* data, the PGC3SCZ summary statistics, and the TGP reference panel (7469969 SNPs). These SNPs were further subset for those that had an imputation INFO score greater than 0.8 in both Bio*Me* and PGC3SCZ, requiring that the difference in INFO scores between Bio*Me* and PGC3SCZ be less than 0.25 (leaving 6,652,763 SNPs). Next, the TGP reference containing all 2,504 individuals was then subset for these 6,652,763 sites and stratified into three datasets, one for the AFR, EUR and AMR ancestral cohorts in TGP, respectively. These three TGP datasets were then separately filtered to remove SNPs that within the given ancestry demonstrating genotype missingness greater than 0.02, minor allele frequency less than 0.01, or deviation from Hardy-Weinberg equilibrium (defined as p-value < 5E-16). Using these three TGP datasets as ancestry-specific references, linkage disequilibrium (LD)-based “clumping” was then performed on the PGC3SCZ summary statistics in order to obtain approximately independent sets of SNPs to be used for PRS calculations in the Bio*Me* subset of the corresponding ancestry. The clumping procedure was carried out three times (one for each of the EUR, AFR and AMR TGP reference datasets prepared) using the “--clump” function in PLINK (v1.90b6.10). For each ancestry, only the SNPs remaining in the TGP reference after filtering were included in the clumping procedure, and clumps were formed for all SNPs within 1000 kilobases of one another (or 10,000 kilobases for chromosomes 6, 8, and 17) found to be in linkage disequilibrium (r^2^ < 0.1), resulting in 302,332 SNPs in AFR, 187,044 SNPs in AMR, and 192,307 SNPs in EUR retained for the PRS calculations. The Bio*Me* imputed dosage data was then stratified into EUR, AMR, and AFR cohorts and subset for the corresponding ancestry-specific set of clumped PGC3SCZ SNPs. PRS were then calculated using SNPs with a PGC3SCZ GWAS p-value below 0.5 with PRSice (v2.2.6)^41^. We evaluated the significance of SCZ*−*control PRS differences within-ancestry using logistic regression including as covariates the top 10 PCs. Predictive values of these scores are reported in terms of Nagelkerke’s pseudo-R^2^, which was calculated in R 3.5.3 using the rms package^42^.

### Genomic Psychiatry Cohort (GPC)

The Genomic Psychiatry Cohort (GPC) is a large multi-ethnic sample of cases with psychotic illness (schizophrenia, schizoaffective disorder, and bipolar disorder) and screened controls^21^. Using a combination of focused, direct interviews and data extraction from medical records, diagnoses were established using the OPCRIT system^22^. For the present analysis, we included individuals with a diagnosis of schizophrenia or schizoaffective disorder. Details of ascertainment, diagnosis, genotyping, quality control, and polygenic risk scoring for the GPC cases included in this report have been previously described^23^. In brief, all participants enrolled as probable cases were interviewed by mental health professionals using the Diagnostic Interview for Psychosis and Affective Disorders (DI-PAD), a semi-structured clinical interview developed specifically for the GPC that is administered. Inclusion criteria for cases include meeting lifetime diagnostic criteria for schizophrenia or schizoaffective disorder (any subtype) in accordance with the OPCRIT algorithms for DSM-IV and/or ICD-10 criteria, and/or DSM-5. Exclusion criteria included any premorbid organic mental disorders (i.e., epilepsy, CNS infection, significant head trauma, mental retardation), and premorbid history of significant drug or alcohol dependence by DSM IV/5 that confounds the diagnosis of schizophrenia. All participants gave written informed consent and the IRB of the participating institutions approved the protocol.

### Variance explained by polygenic risk scores

We calculated the proportion of variance explained for SCZ compared to controls (in Bio*Me*) and for SCZ outcomes (in Bio*Me* and GPC). Nagelkerke’s pseudo-R^2^ was calculated by subtracting the Nagelkerke’s R^2^ attributable to ancestry covariates alone from the Nagelkerke’s R^2^ for PRS plus covariates. These calculations were done in R using the base lm() function and the lrm() function in the rms package.

### Predictive Modeling

Separately, for both Bio*Me* and GPC cohorts, we first created different datasets considering all available features, both genetic and non-genetic, and separate outcomes. Within each dataset, we dropped individuals with missing outcome information and predictors with >70% missing information. For model generalization purposes, we split the datasets into training (70%) and test (30%) sets, stratifying for outcome and ancestry. Within each ancestry, missing values were imputed using median for continuous features and mode for categorical features from the training sets. Continuous features were rescaled between 0 and 1 within each ancestry subgroup. Finally, for each outcome variable, we created three separate datasets including (a) clinical features, (b) PCs and PRS, and (c) both clinical and genetic features, respectively.

Because of class imbalance in Bio*Me* (***Supplementary Table 6***), we binarized the outcome “number of psychiatric admissions” considering as two separate classes the individuals with no psychiatric admissions, and those with at least one psychiatric admission (see ***Table 1***). Both Bio*Me* predictors that reported a significant association are categorical variables and no features were dropped based on the percentage of missing information. ***Table 1*** reports the description of the Bio*Me* dataset and ***Supplementary Table 5*** shows the dataset description broken down by ancestry. To improve the models’ performance, we randomly oversampled the minority class for the internal training datasets within cross validation, with the ratio of the number of samples in the minority class over the number of samples in the majority class after resampling equal to 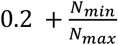, where *N*_*min*_ is the minority class cardinality and *N*_*max*_ the majority class cardinality, before resampling. Dataset descriptions broken down by ancestry can be found in ***Supplementary Table 7***.

For GPC cohort, we dropped 11 features due to >70% missing information. Because of class imbalances we binarized the outcome describing the course of the disorder separating individuals according to their symptoms, i.e., with or without deterioration. ***Table 1*** includes the description of the GPC dataset with outcome class counts. As can be observed, classes are relatively balanced, hence no oversampling methods were used. Dataset descriptions broken down by ancestry can be found in ***Supplementary Table 5***.

To investigate the predictive performance of different sets of features, we fit logistic regression models with *L*_2_ penalty to each dataset and tune the regularization parameter within a 100×3 repeated cross-validation framework (stratified for outcome and ancestry). The regularization parameter varied from 0.001 to 100, with higher values indicating less regularization (see ***Supplementary Figure 1***). To best deal with imbalanced classes, parameter configuration selection during training was done maximizing the *F*_2_ score. The model selected was then trained on the entire training set and evaluated on the test set and the *F*_2_ score was reported as the generalization metric. *F*_2_ score is a general version of the traditional *F*_1_ score defined as 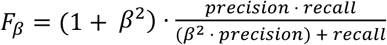, with *β* = *2*, so that recall is weighted higher than precision. Bootstrap estimates of area under the precision-recall curve (AUPRC) were calculated within 100 repetitions to investigate the metrics trade-off at different probability thresholds and determine the best feature configuration in predicting clinical outcomes. To assess the predictive performance of the genetic features compared to clinical features we performed one-way repeated-measure ANOVAs to compare the average in-sample *F*_2_ scores during cross-validation and bootstrapped AUPRCs among different feature configurations. Post-hoc testing for significant results was done via two-sided pairwise t-tests with Benjamini-Hochberg correction (see ***Supplementary Table 9***). This approach is repeated considering subsets of patients with different ancestries among AFR, AMR, and EUR. For these analyses, the stratification was done only considering outcome variables. For datasets with both clinical and genetic, and only genetic features, we also run models with a binarized version of PRS, where all scores greater or equal than the last training set decile are set to 1 and the rest is set to 0. This to leverage more the extreme PRS values and investigate if they could improve the outcome prediction.

All computations were run on a server with Intel Xeon Platinum 8168 2.70GHz 64-bit 24-core processor. Data preprocessing, modeling, and testing were implemented in Python 3.8 using *scikit-learn* library^43^ for machine learning modeling and R 3.5.3 with *boot* library^44,45^ for bootstrap estimates. Code can be found at https://github.mountsinai.org/isotta-landi/prs_schizophrenia.

## Data Availability

Polygenic risk scores and a limited (de-identified) version of the clinical data utilized in this manuscript are available upon request.

## Code availability statement

All code used in the generation of this manuscript is available upon request or can be found at https://github.mountsinai.org/isotta-landi/prs_schizophrenia.

