## Supplementary Material for "Polygenic risk scores lack prognostic value for adults with severe mental illness"

**Tables**

| **Variable Analysis Status** | **Variable Source** | **Variable** | **Variable Description** | **Corresponding GPC Variable** |
| --- | --- | --- | --- | --- |
| Case status | Structured EMR data | SCZ or related diagnosis | Billed for treatment of SCZ or related diagnosis on >=1 occasion | NA |
| Outcome | NLP of clinical notes | Self-harm risk | Characterized by treating clinician as being at risk for self harm in >=1 clinical note | OPCRIT 43 |
| Outcome | NLP of clinical notes | Homelessness | Noted by treating clinician to be undomiciled in >=1 clinical note | NA |
| Outcome | NLP of clinical notes | Aggressive behavior | Episode(s) of aggressive behavior recorded by treating clinician in >=1 clinical note | NA |
| Outcome | Structured EMR data | History of clozapine prescription | Prescribed clozapine in MSHS on >=1 occasion | NA |
| Outcome | Structured EMR data | Number of unique antipsychotics prescribed | The number of unique antipsychotics prescribed to patient in their history of treatment within the MSHS (binarized to <2 vs. >=2) | NA |
| Outcome | Structured EMR data | Number of psychiatric hospitalizations | The number of times the patient was admitted to a psychiatric ward within the MSHS (binarized to 0 vs. greater than 0) | NA |
| Outcome predictor | Bio*Me* questionnaire | Family History of SCZ (grandparents or first degree relative) | Binary self-reported indicator of SCZ family history | OPCRIT 13 |
| Outcome predictor | Bio*Me* questionnaire | Family History of depression (grandparents or first degree relative) | Binary self-reported indicator of depression family history | OPCRIT 14 |
| Outcome predictor | Bio*Me* questionnaire | Family History of autism (grandparents or first degree relative) | Binary self-reported indicator of autism family history | OPCRIT 14 |
| Outcome predictor | Bio*Me* questionnaire | Adoption history | Binary self-reported indicator of whether the patient was adopted | NA |
| Outcome predictor | Bio*Me* questionnaire | Smoking history | Binary self-reported indicator of whether the patient has ever been a cigarette smoker | NA |
| Outcome predictor | Bio*Me* questionnaire | Migration history | Binary self-reported indicator of whether the patient migrated to the United States from another country | NA |
| Outcome predictor | NLP of clinical notes | Delusions | Delusions noted by treating clinician in >=1 clinical note | DIPAD screen for delusions |
| Outcome predictor | NLP of clinical notes | Bizarre behavior | Bizarre behavior noted by treating clinician in >=1 clinical note | OPCRIT 17 |
| Outcome predictor | NLP of clinical notes | Decreased need for sleep | Decreased need for sleep noted by treating clinician in >=1 clinical note | OPCRIT 22 |
| Outcome predictor | NLP of clinical notes | Shouting | Shouting noted by treating clinician in >=1 clinical note | OPCRIT 23 |
| Outcome predictor | NLP of clinical notes | Motor retardation | Motor retardation noted by treating clinician in >=1 clinical note | OPCRIT 24 |
| Outcome predictor | NLP of clinical notes | Incoherent thinking | Incoherent thinking noted by treating clinician in >=1 clinical note | OPCRIT 27 |
| Outcome predictor | NLP of clinical notes | Pressured speech | Pressured speech noted by treating clinician in >=1 clinical note | OPCRIT 30 |
| Outcome predictor | NLP of clinical notes | Racing thoughts | Racing thoughts noted by treating clinician in >=1 clinical note | OPCRIT 31 |
| Outcome predictor | NLP of clinical notes | Blunted affect | Blunted affect noted by treating clinician in >=1 clinical note | OPCRIT 33 |
| Outcome predictor | NLP of clinical notes | Inappropriate affect | Inappropriate affect noted by treating clinician in >=1 clinical note | OPCRIT 34 |
| Outcome predictor | NLP of clinical notes | Elevated mood | Elevated mood noted by treating clinician in >=1 clinical note | OPCRIT 35 |
| Outcome predictor | NLP of clinical notes | Irritable mood | Irritable mood noted by treating clinician in >=1 clinical note | OPCRIT 36 |
| Outcome predictor | NLP of clinical notes | Indifference | Indifference noted by treating clinician in >=1 clinical note | OPCRIT 39 |
| Outcome predictor | NLP of clinical notes | Guilt | Guilt noted by treating clinician in >=1 clinical note | OPCRIT 42 |
| Outcome predictor | NLP of clinical notes | Inflated opinion of self | Inflated opinion of self noted by treating clinician in >=1 clinical note | OPCRIT 56 |
| Outcome predictor | NLP of clinical notes | Hallucinations | Hallucinations noted by treating clinician in >=1 clinical note | OPCRIT 77 |
| Outcome predictor | NLP of clinical notes | Impaired insight | Impaired insight noted by treating clinician in >=1 clinical note | OPCRIT 85 |
| Outcome predictor | NLP of clinical notes | Paranoia | Paranoia noted by treating clinician in >=1 clinical note | OPCRIT 54 |
| Outcome predictor | NLP of clinical notes | Perseveration | Perseveration noted by treating clinician in >=1 clinical note | OPCRIT 28 |
| Outcome predictor | NLP of clinical notes | Circumstantiality | Circumstantiality noted by treating clinician in >=1 clinical note | OPCRIT 28 |
| Outcome predictor | NLP of clinical notes | Ideas of reference | Ideas of reference noted by treating clinician in >=1 clinical note | OPCRIT 58 |
| Outcome predictor | NLP of clinical notes | Tangentiality | Tangentiality noted by treating clinician in >=1 clinical note | OPCRIT 28 |
| Outcome predictor | NLP of clinical notes | Impulsivity | Impulsivity noted by treating clinician in >=1 clinical note | OPCRIT 20 |
| Outcome predictor | NLP of clinical notes | Thought deprivation | Thought deprivation noted by treating clinician in >=1 clinical note | OPCRIT 29/OPCRIT 67 |
| Outcome predictor | NLP of clinical notes | Disruptive behavior | Disruptive behavior noted by treating clinician in >=1 clinical note | OPCRIT 20 |
| Outcome predictor | NLP of clinical notes | Disturbance in structure of associations | Disturbance in structure of associations noted by treating clinician in >=1 clinical note | OPCRIT 28 |
| Outcome predictor | NLP of clinical notes | Inappropriate behavior | Inappropriate behavior noted by treating clinician in >=1 clinical note | OPCRIT 17 |
| Outcome predictor | NLP of clinical notes | Uncooperative behavior | Uncooperative behavior noted by treating clinician in >=1 clinical note | OPCRIT 86 |
| Outcome predictor | NLP of clinical notes | Unkempt appearance | Unkempt appearance noted by treating clinician in >=1 clinical note | NA |
| Outcome predictor | Bio*Me* questionnaire | Gender | Self-reported gender | OPCRIT 03 |
| Outcome predictor | Structured EMR data | Relationship Status | Marital status recorded in EMR | OPCRIT 06 |
| Outcome predictor | Structured EMR data | Trauma-related comorbidities | Billed for treatment of trauma-related psychiatric condition on >=1 occasion | NA |
| Outcome predictor | Structured EMR data | Substance-related comorbidities | Billed for treatment of substance-related psychiatric condition on >=1 occasion | NA |

**Supplementary Table 1. Case definition, outcomes, and clinical features for Bio*Me* dataset.**

| **DIPAD Section** | **Variable Group** | **Variable** | **Variable Analysis Status** | **Variable Description** | **Variable Class** | **Variable Values** |
| --- | --- | --- | --- | --- | --- | --- |
| Participant Ratings | Details and history | Age | Outcome predictor | Age at interview | Continuous | NA |
| Participant Ratings | Abnormal beliefs and ideas | Delusions | Outcome predictor | Screen for delusions | Binary categorical | No delusions\|Delusions present |
| Participant Ratings | Details and history | OPCRIT 01 | Outcome predictor | Source of rating | Categorical | Hospital case notes (charts)\|Structured interview with subject\|Prepared abstract\|Interview with informant\|Combined sources including structured interview\|Combined sources not including structured interview |
| Participant Ratings | Details and history | OPCRIT 03 | Outcome predictor | Gender | Binary categorical | Female\|Male |
| Participant Ratings | Details and history | OPCRIT 04 | Outcome predictor | Age of onset | Continuous | NA |
| Participant Ratings | Details and history | OPCRIT 05 | Outcome predictor | Mode of onset | Ordered categorical | No episode\|Abrupt onset definable within hours or days\|Acute onset definable within one month\|Moderately acute onset definable within one month\|Gradual onset over period up to six months\|Insidious onset over period greater than six months |
| Participant Ratings | Details and history | OPCRIT 06 | Outcome predictor | Single | Binary categorical | Married; Has been married or has lived with the same partner for at least 6 months\|Single; has never married or lived as married |
| Participant Ratings | Details and history | OPCRIT 07 | Outcome predictor | Employment status at onset | Binary categorical | Employed at onset (includes full time students and homemakers)\|Not employed at onset |
| Interviewer Ratings | Details and history | OPCRIT 08 | Excluded (missingness) | Duration of illness in weeks | Numeric | NA |
| Participant Ratings | Details and history | OPCRIT 09 | Outcome predictor | Premorbid work adjustment | Binary categorical | Good premorbid work adjustment\|Poor premorbid work adjustment |
| Participant Ratings | Details and history | OPCRIT 10 | Outcome predictor | Premorbid social adjustment | Binary categorical | Good premorbid social adjustment\|Poor premorbid social adjustment |
| Participant Ratings | Details and history | OPCRIT 13 | Outcome predictor | Family history of schizophrenia | Binary categorical | No family history of schizophrenia\|Family history of schizophrenia |
| Participant Ratings | Details and history | OPCRIT 14 | Outcome predictor | Family history of psychiatric disorder other than schizophrenia | Binary categorical | No family history\|Family history of psychiatric disorder other than schizophrenia |
| Participant Ratings | Details and history | OPCRIT 15 | Outcome predictor | Coarse brain disorder prior to onset | Binary categorical | No prior brain disease present\|Prior brain disease present |
| Participant Ratings | Details and history | OPCRIT 16 | Outcome predictor | Psychosocial stressors prior to onset of first episode | Binary categorical | Stressor not present\|Stressor present |
| Interviewer Ratings | Appearance and behavior | OPCRIT 17 | Outcome predictor | Bizarre behaviour | Binary categorical | Not present\|Present |
| Interviewer Ratings | Appearance and behavior | OPCRIT 18 | Outcome predictor | Catatonia | Ordered categorical | Not present\|Present less than one month\|Present most of the time in a one month period or longer |
| Participant Ratings | Appearance and behavior | OPCRIT 19 | Excluded (missingness) | Excessive activity | Ordered categorical | Not present\|Present at least four days\|Present at least one week\|Present at least two weeks |
| Participant Ratings | Appearance and behavior | OPCRIT 20 | Excluded (missingness) | Reckless activity | Ordered categorical | Not present\|Present at least four days\|Present at least one week\|Present at least two weeks |
| Participant Ratings | Appearance and behavior | OPCRIT 21 | Excluded (missingness) | Distractibility | Ordered categorical | Not present\|Present at least four days\|Present at least one week\|Present at least two weeks |
| Participant Ratings | Appearance and behavior | OPCRIT 22 | Excluded (missingness) | Reduced need for sleep | Ordered categorical | Not present\|Present at least four days\|Present at least one week\|Present at least two weeks |
| Participant Ratings | Appearance and behavior | OPCRIT 22a | Excluded (missingness) | Reduced need for sleep number hours normally sleep | Continuous | NA |
| Participant Ratings | Appearance and behavior | OPCRIT 22b | Excluded (missingness) | Reduced need for sleep number hours sleeping during reduced period | Continuous | NA |
| Interviewer Ratings | Appearance and behavior | OPCRIT 23 | Outcome predictor | Agitated activity | Ordered categorical | Not present\|Present at least one week\|Present at least two weeks\|Present at least one month |
| Participant Ratings | Appearance and behavior | OPCRIT 24 | Outcome predictor | Slowed activity | Ordered categorical | Not Present;Present at least one week;Present at least two weeks;Present at least one month\|Not Present;Present at least one week;Present at least two weeks;Present at least one month\|Not Present;Present at least one week;Present at least two weeks;Present at least one month\|Not Present;Present at least one week;Present at least two weeks;Present at least one month |
| Participant Ratings | Appearance and behavior | OPCRIT 25 | Outcome predictor | Loss of energy or fatigue | Ordered categorical | Not Present;Present at least one week;Present at least two weeks;Present at least one month\|Not Present;Present at least one week;Present at least two weeks;Present at least one month\|Not Present;Present at least one week;Present at least two weeks;Present at least one month\|Not Present;Present at least one week;Present at least two weeks;Present at least one month |
| Interviewer Ratings | Speech and form of thought | OPCRIT 26 | Outcome predictor | Speech difficult to understand | Binary categorical | Not present\|Present |
| Interviewer Ratings | Speech and form of thought | OPCRIT 27 | Outcome predictor | Incoherent | Ordered categorical | Not present\|Present less than one month\|Present most of the time in a one month period or longer |
| Interviewer Ratings | Speech and form of thought | OPCRIT 28 | Outcome predictor | Positive formal thought disorder | Ordered categorical | Not present\|Present less than one month\|Present most of the time in a one month period or longer |
| Interviewer Ratings | Speech and form of thought | OPCRIT 29 | Outcome predictor | Negative formal thought disorder | Ordered categorical | Not present\|Present less than one month\|Present most of the time in a one month period or longer |
| Participant Ratings | Speech and form of thought | OPCRIT 30 | Excluded (missingness) | Pressured speech | Ordered categorical | Not present\|Present at least four days\|Present at least one week\|Present at least two weeks |
| Participant Ratings | Speech and form of thought | OPCRIT 31 | Excluded (missingness) | Racing thoughts | Ordered categorical | Not present\|Present at least four days\|Present at least one week\|Present at least two weeks |
| Interviewer Ratings | Affect and associated features | OPCRIT 32 | Outcome predictor | Restricted affect | Ordered categorical | Not present\|Present less than one month\|Present most of the time in a one month period or longer |
| Interviewer Ratings | Affect and associated features | OPCRIT 33 | Outcome predictor | Blunted affect | Ordered categorical | Not present\|Present less than one month\|Present most of the time in a one month period or longer |
| Interviewer Ratings | Affect and associated features | OPCRIT 34 | Outcome predictor | Inappropriate affect | Ordered categorical | Not present\|Present less than one month\|Present most of the time in a one month period or longer |
| Participant Ratings | Affect and associated features | OPCRIT 35 | Outcome predictor | Elevated mood | Ordered categorical | Not present\|Present at least four days\|Present at least one week\|Present at least two weeks OR if lasted < one week but hospitalized for affective disorder |
| Participant Ratings | Affect and associated features | OPCRIT 36 | Outcome predictor | Irritable mood | Ordered categorical | Not present\|Present at least four days\|Present at least one week\|Present at least two weeks OR if lasted < one week but hospitalized for affective disorder |
| Participant Ratings | Affect and associated features | OPCRIT 36a | Outcome predictor | Number of manic episodes | Continuous | NA |
| Participant Ratings | Affect and associated features | OPCRIT 37 | Outcome predictor | Length of most severe episode of either depressed or irritible mood | Ordered categorical | Not present\|Present at least one week\|Present at least two weeks\|Present at least one month |
| Participant Ratings | Affect and associated features | OPCRIT 37a | Outcome predictor | Depressed for two weeks | Binary categorical | Not Present\|Present |
| Participant Ratings | Affect and associated features | OPCRIT 37b | Outcome predictor | Irritible for one week | Binary categorical | Not present\|Present |
| Participant Ratings | Affect and associated features | OPCRIT 37d | Outcome predictor | Number of depressive episodes | Continuous | NA |
| Participant Ratings | Affect and associated features | OPCRIT 38 | Outcome predictor | Diurnal variation | Binary categorical | No depression, or not worse early\|Regularly feels worse early in the day |
| Participant Ratings | Affect and associated features | OPCRIT 39 | Outcome predictor | Loss of pleasure | Ordered categorical | Not Present;Present at least one week;Present at least two weeks;Present at least one month\|Not Present;Present at least one week;Present at least two weeks;Present at least one month\|Not Present;Present at least one week;Present at least two weeks;Present at least one month\|Not Present;Present at least one week;Present at least two weeks;Present at least one month |
| Participant Ratings | Affect and associated features | OPCRIT 40 | Outcome predictor | Altered libido | Categorical | No change\|Loss of libido for at least one week\|Increase in libido for at least one week |
| Participant Ratings | Affect and associated features | OPCRIT 41 | Outcome predictor | Impaired concentration | Ordered categorical | Not Present;Present at least one week;Present at least two weeks;Present at least one month\|Not Present;Present at least one week;Present at least two weeks;Present at least one month\|Not Present;Present at least one week;Present at least two weeks;Present at least one month\|Not Present;Present at least one week;Present at least two weeks;Present at least one month |
| Participant Ratings | Affect and associated features | OPCRIT 42 | Outcome predictor | Excessive self reproach | Ordered categorical | Not present\|Present at least one week\|Present at least two weeks\|Present at least one month |
| Participant Ratings | Affect and associated features | OPCRIT 43 | Outcome | Suicidality | Ordered categorical | Not present\|Suicidal ideation present at least one week\|Suicidal ideation present at least two weeks or suicide attempt\|Suicidal ideation present at least one month or suicide attempt |
| Participant Ratings | Affect and associated features | OPCRIT 44 | Outcome predictor | Initial insomnia | Ordered categorical | Not present\|Present at least one week\|Present at least two weeks\|Present at least one month |
| Participant Ratings | Affect and associated features | OPCRIT 45 | Outcome predictor | Middle insomnia | Binary categorical | No waking\|Middle insomnia present |
| Participant Ratings | Affect and associated features | OPCRIT 46 | Outcome predictor | Terminal insomnia | Ordered categorical | Not present\|Present at least one week\|Present at least two weeks\|Present at least one month |
| Participant Ratings | Affect and associated features | OPCRIT 47 | Outcome predictor | Excessive sleep | Ordered categorical | Not present\|Present at least one week\|Present at least two weeks\|Present at least one month |
| Participant Ratings | Affect and associated features | OPCRIT 47a | Outcome predictor | Excessive sleep number hours normally sleep | Continuous | NA |
| Participant Ratings | Affect and associated features | OPCRIT 47b | Outcome predictor | Excessive sleep number hours sleeping during excessive period | Continuous | NA |
| Participant Ratings | Affect and associated features | OPCRIT 48 | Outcome predictor | Decreased appetite | Ordered categorical | Not present\|Present at least one week\|Present at least two weeks\|Present at least one month |
| Participant Ratings | Affect and associated features | OPCRIT 49 | Outcome predictor | Weight loss | Ordered categorical | No loss\|Loss of at least 1+ lb (0.5 kg) per week for 3 or more weeks - or - 4 lbs (2 kg) in less than 3 weeks\|Loss of at least 2+ lbs (1 kg) per week for 3 or more weeks - or - 5 to 9 lbs (2.5 to 4.5 kg) in less than 3 weeks\|Loss of at least 10+ lbs (5 kg) in less than 1 year |
| Participant Ratings | Affect and associated features | OPCRIT 50 | Outcome predictor | Increased appetite | Ordered categorical | Not present\|Present at least one week\|Present at least two weeks\|Present at least one month |
| Participant Ratings | Affect and associated features | OPCRIT 51 | Outcome predictor | Weight gain | Ordered categorical | No Gain\|Gain of at least 1+ lb (0.5 kg) per week for 3 or more week - or - 4 lbs (2 kg) in less than 3 weeks\|Gain of at least 2+ lbs (1 kg) per week for 3 or more weeks - or - 5 to 9 lbs (2.5 to 4.5 kg) in less than 3 weeks\|Gain of at least 10+ lbs (5 kg) in less than 1 year |
| Participant Ratings | Affect and associated features | OPCRIT 52 | Outcome predictor | Relationship between psychotic and affective symptoms | Categorical | No co-occurrence\|Psychotic symptoms dominate the clinical picture although occasional affective disturbance may also occur.\|Psychotic and affective symptoms are balanced, with neither group of symptoms dominating the overall course of the illness.\|Affective symptoms predominate although psychotic symptoms may also occur. |
| Participant Ratings | Affect and associated features | OPCRIT 53 | Excluded (missingness) | Increased sociability | Ordered categorical | Not present\|Present at least four days\|Present at least one week\|Present at least two weeks |
| Participant Ratings | Abnormal beliefs and ideas | OPCRIT 54 | Outcome predictor | Persecutory delusions | Ordered categorical | Not present\|Present less than one month\|Present most of the time in a one month period or longer |
| Interviewer Ratings | Abnormal beliefs and ideas | OPCRIT 55 | Outcome predictor | Well organised delusions | Ordered categorical | Not present\|Present less than one month\|Present most of the time in a one month period or longer |
| Participant Ratings | Abnormal beliefs and ideas | OPCRIT 56 | Excluded (missingness) | Increased self esteem | Ordered categorical | Not present\|Present at least four days\|Present at least one week\|Present at least two weeks |
| Participant Ratings | Abnormal beliefs and ideas | OPCRIT 57 | Outcome predictor | Grandiose delusions | Ordered categorical | Not present\|Present at least four days\|Present at least one week\|Present at least two weeks |
| Participant Ratings | Abnormal beliefs and ideas | OPCRIT 58 | Outcome predictor | Delusions of influence | Ordered categorical | Not present\|Present less than one month\|Present most of the time in a one month period or longer |
| Participant Ratings | Abnormal beliefs and ideas | OPCRIT 59 | Outcome predictor | Bizarre delusions | Ordered categorical | Not present\|Present less than one month\|Present most of the time in a one month period or longer |
| Interviewer Ratings | Abnormal beliefs and ideas | OPCRIT 60 | Outcome predictor | Widespread delusions | Ordered categorical | Not present\|Present less than one month\|Present most of the time in a one month period or longer |
| Participant Ratings | Abnormal beliefs and ideas | OPCRIT 61 | Outcome predictor | Delusions of passivity | Ordered categorical | Not present\|Present less than one month\|Present most of the time in a one month period or longer |
| Participant Ratings | Abnormal beliefs and ideas | OPCRIT 62 | Outcome predictor | Primary delusional ideas | Ordered categorical | Not present\|Present less than one month\|Present most of the time in a one month period or longer |
| Participant Ratings | Abnormal beliefs and ideas | OPCRIT 63 | Outcome predictor | Delusional moods | Ordered categorical | Not present\|Present less than one month\|Present most of the time in a one month period or longer |
| Interviewer Ratings | Abnormal beliefs and ideas | OPCRIT 64 | Outcome predictor | Delusions and hallucinations last for one week | Ordered categorical | Not present\|Present less than one month\|Present most of the time in a one month period or longer |
| Interviewer Ratings | Abnormal beliefs and ideas | OPCRIT 65 | Outcome predictor | Persecutory or jealous delusions and hallucinations | Ordered categorical | Not present\|Present less than one month\|Present most of the time in a one month period or longer |
| Participant Ratings | Abnormal beliefs and ideas | OPCRIT 66 | Outcome predictor | Thought insertion | Ordered categorical | Not present\|Present less than one month\|Present most of the time in a one month period or longer |
| Participant Ratings | Abnormal beliefs and ideas | OPCRIT 67 | Outcome predictor | Thought withdrawal | Ordered categorical | Not present\|Present less than one month\|Present most of the time in a one month period or longer |
| Participant Ratings | Abnormal beliefs and ideas | OPCRIT 68 | Outcome predictor | Thought broadcast | Ordered categorical | Not present\|Present less than one month\|Present most of the time in a one month period or longer |
| Participant Ratings | Abnormal beliefs and ideas | OPCRIT 69 | Outcome predictor | Delusions of guilt | Ordered categorical | No delusions of guilt\|Present at least one week\|Present at least two weeks\|Present at least one month |
| Participant Ratings | Abnormal beliefs and ideas | OPCRIT 70 | Outcome predictor | Delusions of poverty | Ordered categorical | No delusions of poverty\|Present at least one week\|Present at least two weeks\|Present at least one month |
| Participant Ratings | Abnormal beliefs and ideas | OPCRIT 71 | Outcome predictor | Nihilistic delusions | Ordered categorical | No nihilistic delusions\|Present at least one week\|Present at least two weeks\|Present at least one month |
| Participant Ratings | Abnormal perceptions | OPCRIT 72 | Outcome predictor | Thought echo | Ordered categorical | Not present\|Present less than one month\|Present most of the time in a one month period or longer |
| Participant Ratings | Abnormal perceptions | OPCRIT 73 | Outcome predictor | Third person auditory hallucinations | Ordered categorical | Not present\|Present less than one month\|Present most of the time in a one month period or longer |
| Participant Ratings | Abnormal perceptions | OPCRIT 74 | Outcome predictor | Running commentary voices | Ordered categorical | Not present\|Present less than one month\|Present most of the time in a one month period or longer |
| Participant Ratings | Abnormal perceptions | OPCRIT 75 | Outcome predictor | Command accusatory abusive or persecutory voices | Ordered categorical | Not present\|Present less than one month\|Present most of the time in a one month period or longer |
| Participant Ratings | Abnormal perceptions | OPCRIT 76 | Outcome predictor | Neutral or nonverbal hallucinations | Ordered categorical | Not present\|Present less than one month\|Present most of the time in a one month period or longer |
| Participant Ratings | Abnormal perceptions | OPCRIT 77 | Outcome predictor | Hallucination in any modality | Ordered categorical | Not present\|Present less than one month\|Present most of the time in a one month period or longer |
| Interviewer Ratings | General appraisal | OPCRIT 84 | Outcome predictor | Information not credible | Binary categorical | Information credible\|Information not credible |
| Participant Ratings | General appraisal | OPCRIT 85 | Outcome predictor | Insight | Binary categorical | Insight present\|Insight impaired |
| Interviewer Ratings | General appraisal | OPCRIT 86 | Outcome predictor | Rapport difficult | Binary categorical | Rapport established without difficulty\|Rapport difficult |
| Interviewer Ratings | General appraisal | OPCRIT 87 | Excluded (no variation) | Impairment or incapacity during disorder | Ordered categorical | No impairment.\|Subjective impairment only (at work, school, family or social functioning).\|Evidence of objective functional impairment in major life role with definite reduction in productivity and/or criticism about functioning or performance has been received.\|Inpatient treatment (any duration) has been received, or no functioning at all in major life role for more than two days, or active psychotic symptoms such as delusions or hallucinations have occurred. |
| Interviewer Ratings | General appraisal | OPCRIT 88 | Excluded (no variation) | Deterioration from premorbid level of functioning | Binary categorical | No deterioration present\|Deterioration from premorbid level of functioning |
| Participant Ratings | General appraisal | OPCRIT 89 | Excluded (no variation) | Psychotic symptoms respond to antipsychotic medications | Binary categorical | No response to neuroleptics, never been psychotic, or never taken neuroleptics\|Positive response to neuroleptics |
| Interviewer Ratings | General appraisal | OPCRIT 90 | Outcome | Course of disorder | Binary categorical | Continuous chronic illness\|Continuous chronic illness with deterioration |
| Participant Ratings | Affect and associated features | Schizoaffective | Outcome predictor | Psychotic symptoms for 2 weeks without mood symptoms | Binary categorical | Yes\|No |

**Supplementary Table 2. Case definition, outcomes, and clinical features for Genomic Psychiatry Cohort (GPC) dataset.**

| **Cohort** | **Variable** | **Ancestry** | **r2** | **Beta** | **Standard Error** | **T** | **p-value** |
| --- | --- | --- | --- | --- | --- | --- | --- |
| Bio*Me* | Case Status | ALL | 0.01 | 0.06 | 0.01 | 6.04740698 | 1.49E-09 |
| Bio*Me* | Case Status | EUR | 0.02 | 0.05 | 0.01 | 4.94564448 | 7.71E-07 |
| Bio*Me* | Case Status | AMR | 0.01 | 0.14 | 0.03 | 5.19339164 | 2.11E-07 |
| Bio*Me* | Case Status | AFR | 0.01 | 0.18 | 0.04 | 4.43009802 | 9.56E-06 |
| Bio*Me* | Aggressive Behavior | ALL | 0.03 | 0.36 | 0.12 | 2.96002893 | 0.00317293 |
| Bio*Me* | Aggressive Behavior | EUR | 0.06 | -0.16 | 0.28 | -0.5737458 | 0.56785645 |
| Bio*Me* | Aggressive Behavior | AMR | 0.04 | 0.80 | 0.28 | 2.88584535 | 0.00419272 |
| Bio*Me* | Aggressive Behavior | AFR | 0.05 | 0.76 | 0.28 | 2.74572181 | 0.0063718 |
| Bio*Me* | History of Clozapine Prescription | ALL | 0.00 | 0.07 | 0.12 | 0.58561778 | 0.55830833 |
| Bio*Me* | History of Clozapine Prescription | EUR | 0.02 | 0.46 | 0.43 | 1.07416341 | 0.28619322 |
| Bio*Me* | History of Clozapine Prescription | AMR | 0.00 | 0.13 | 0.22 | 0.58250325 | 0.56067422 |
| Bio*Me* | History of Clozapine Prescription | AFR | 0.01 | -0.29 | 0.26 | -1.1117335 | 0.26706944 |
| Bio*Me* | Number of Unique Antipsychotics Prescribed | ALL | 0.01 | 0.42 | 0.19 | 2.20732359 | 0.02759313 |
| Bio*Me* | Number of Unique Antipsychotics Prescribed | EUR | 0.01 | 0.58 | 0.52 | 1.12445562 | 0.26440578 |
| Bio*Me* | Number of Unique Antipsychotics Prescribed | AMR | 0.00 | 0.44 | 0.41 | 1.07017083 | 0.28542021 |
| Bio*Me* | Number of Unique Antipsychotics Prescribed | AFR | 0.00 | 0.10 | 0.44 | 0.23270465 | 0.81613634 |
| Bio*Me* | Number of Psychiatric Admissions | ALL | 0.02 | 0.52 | 0.18 | 2.89222665 | 0.0039358 |
| Bio*Me* | Number of Psychiatric Admissions | EUR | 0.00 | -0.18 | 0.47 | -0.3819343 | 0.70359012 |
| Bio*Me* | Number of Psychiatric Admissions | AMR | 0.02374973 | 0.93301823 | 0.39663338 | 2.35234417 | 0.01931475 |
| Bio*Me* | Number of Psychiatric Admissions | AFR | 0.0070748 | 0.53415283 | 0.41605351 | 1.2838561 | 0.20010126 |
| Bio*Me* | History of Homelessness | ALL | 0.00381921 | 0.09056548 | 0.10099555 | 0.89672742 | 0.37015174 |
| Bio*Me* | History of Homelessness | EUR | 0.00095142 | 0.56870022 | 0.24068757 | 2.36281506 | 0.0207305 |
| Bio*Me* | History of Homelessness | AMR | -0.002015 | -0.092375 | 0.19579166 | -0.4718026 | 0.63741765 |
| Bio*Me* | History of Homelessness | AFR | 0.01641087 | 0.4133244 | 0.26238492 | 1.57525974 | 0.11616345 |
| Bio*Me* | At Risk for Self Harm | ALL | 0.01531341 | 0.15222582 | 0.08131131 | 1.87213592 | 0.06157622 |
| Bio*Me* | At Risk for Self Harm | EUR | 2.86E-05 | 0.03691987 | 0.18657325 | 0.19788407 | 0.84367113 |
| Bio*Me* | At Risk for Self Harm | AMR | 0.01596046 | 0.28233617 | 0.18951806 | 1.4897587 | 0.13735967 |
| Bio*Me* | At Risk for Self Harm | AFR | 0.00326112 | 0.05787671 | 0.18999175 | 0.3046275 | 0.76084365 |
| GPC | Course of Disorder (OPCRIT 90) | ALL | 0.00165761 | 0.1809978 | 0.05779873 | 3.13151851 | 0.00174572 |
| GPC | Course of Disorder (OPCRIT 90) | AFR | 2.43E-05 | 0.04507947 | 0.18264553 | 0.24681397 | 0.80506709 |
| GPC | Course of Disorder (OPCRIT 90) | AMR | 0.00079069 | 0.11620677 | 0.19730911 | 0.58895794 | 0.55612698 |
| GPC | Course of Disorder (OPCRIT 90) | EUR | 0.007845 | 0.48420657 | 0.10516761 | 4.60414146 | 4.29E-06 |
| GPC | Suicidality (OPCRIT 43) | ALL | 0.0001685 | -0.1867798 | 0.16110153 | -1.159392 | 0.24635377 |
| GPC | Suicidality (OPCRIT 43) | AFR | 6.51E-05 | -0.2039227 | 0.53108216 | -0.3839758 | 0.70103373 |
| GPC | Suicidality (OPCRIT 43) | AMR | 0.00363462 | 0.6702342 | 0.53346159 | 1.25638699 | 0.2097995 |
| GPC | Suicidality (OPCRIT 43) | EUR | 0.00011925 | -0.2407306 | 0.28795767 | -0.835993 | 0.40324788 |

ALL: entire cohort; AFR: African ancestry; AMR: admixed American ancestry; EUR: European ancestry.

**Supplementary Table 3. Variance explained by the SCZ PRS for case status and outcomes in Bio*Me* and Genomic Psychiatry Cohort (GPC).**

| **ICD Code** | **ICD Description** | **N** | **Fraction with ≥1 other code** | **Mean Unique APs** | **Median Unique APs** | **Mean Total APs** | **Median Total APs** |
| --- | --- | --- | --- | --- | --- | --- | --- |
| F209 | Schizophrenia, unspecified | 409 | 0.72 | 2.40 | 2.00 | 32.19 | 14.00 |
| F29 | Unspecified psychosis not due to a substance or known physiological condition | 307 | 0.56 | 2.42 | 2.00 | 22.68 | 11.00 |
| F200 | Paranoid schizophrenia | 274 | 0.90 | 2.68 | 2.00 | 45.41 | 30.00 |
| F259 | Schizoaffective disorder, unspecified | 268 | 0.82 | 2.91 | 3.00 | 41.85 | 26.50 |
| F22 | Delusional disorders | 149 | 0.40 | 1.73 | 1.00 | 16.01 | 5.00 |
| F203 | Undifferentiated schizophrenia | 115 | 0.98 | 2.63 | 2.00 | 54.50 | 35.00 |
| F250 | Schizoaffective disorder, bipolar type | 104 | 0.90 | 3.27 | 3.00 | 44.47 | 36.50 |
| F251 | Schizoaffective disorder, depressive type | 57 | 0.98 | 3.00 | 3.00 | 55.82 | 51.00 |
| F201 | Disorganized schizophrenia | 52 | 1.00 | 3.06 | 3.00 | 53.12 | 44.50 |
| F2089 | Other schizophrenia | 51 | 1.00 | 2.86 | 2.00 | 55.31 | 46.00 |
| F205 | Residual schizophrenia | 45 | 0.98 | 2.73 | 3.00 | 58.11 | 46.00 |
| F258 | Other schizoaffective disorders | 41 | 0.95 | 3.17 | 3.00 | 46.54 | 41.00 |
| F23 | Brief psychotic disorder | 25 | 0.88 | 2.68 | 2.00 | 31.64 | 24.00 |
| F202 | Catatonic schizophrenia | 17 | 0.94 | 2.82 | 3.00 | 71.29 | 57.00 |
| F2081 | Schizophreniform disorder | 16 | 0.75 | 2.69 | 2.50 | 36.06 | 26.50 |
| F21 | Schizotypal disorder | 12 | 0.67 | 2.00 | 2.50 | 33.58 | 8.50 |
| F28 | Other psychotic disorder not due to a substance or known physiological condition | 8 | 0.75 | 3.25 | 3.00 | 22.13 | 16.50 |
| F24 | Shared psychotic disorder | 1 | 1.00 | 1.00 | 1.00 | 19.00 | 19.00 |

AP: antipsychotic.

**Supplementary Table 4. Summary of diagnostic codes used to define the BioMe case cohort.**

| **Bio*Me* (N=762)** | | | | | |
| --- | --- | --- | --- | --- | --- |
| **Ancestry** | **Gender** | **Outcomes** | **Class counts 0/1** | **N** | **Mean age (sd)** |
| AFR | F | Aggressive | 175/18 | 193 | 60.23 (16.24) |
| AFR | F | Psychiatric admissions | 140/54 |  |  |
| AFR | M | Aggressive | 138/18 | 156 | 59.62 (16.58) |
| AFR | M | Psychiatric admissions | 112/44 |  |  |
| AMR | F | Aggressive | 142/19 | 161 | 58.55 (13.99) |
| AMR | F | Psychiatric admissions | 119/42 |  |  |
| AMR | M | Aggressive | 136/19 | 155 | 60.10 (16.15) |
| AMR | M | Psychiatric admissions | 104/51 |  |  |
| EUR | F | Aggressive | 32/5 | 37 | 53.38 (15.28) |
| EUR | F | Psychiatric admissions | 24/13 |  |  |
| EUR | M | Aggressive | 58/2 | 60 | 60.30 (16.58) |
| EUR | M | Psychiatric admissions | 48/12 |  |  |
| **GPC (N=7,779)** | | | | | |
| **Ancestry** | **Gender** | **Outcomes** | **Class counts 0/1** | **N** | **Mean age (sd)** |
| AFR | F | Course of disorder | 741/420 | 1,161 | 44.25 (11.74) |
| AFR | M | Course of disorder | 1,466/895 | 2,361 | 42.54 (12.61) |
| AMR | F | Course of disorder | 119/99 | 218 | 43.02 (12.36) |
| AMR | M | Course of disorder | 206/193 | 399 | 39.72 (11.91) |
| EUR | F | Course of disorder | 604/609 | 1,213 | 47.1 (12.45) |
| EUR | M | Course of disorder | 1,169/1256 | 2,425 | 44.32 (12.53) |

GPC: Genomic Psychiatry Cohort; AFR: African; AMR: admixed American; EUR: European; F: female; M: male.

**Supplementary Table 5. Description of Bio*Me* and Genomic Psychiatry Cohort (GPC) cohorts by ancestry.**

| **Bio*Me*** | | | | | | |
| --- | --- | --- | --- | --- | --- | --- |
|  |  |  | **Train** | | **Test** | |
| **Outcome** | **Description** | **Max features (clinical + genetic)** | **Samples** | **Class counts (N)** | **Samples** | **Class counts (N)** |
| Aggressive | Development of aggressive behavior | 61 (40 + 21) | 536 | 0 (478); 1 (58) | 226 | 0 (203); 1 (23) |
| Psychiatric admissions | Number of psychiatric admissions | 61 (40 + 21) | 537 | 0 (385); 1 (152) | 225 | 0 (162); 1 (63) |
| **GPC** | | | | | | |
|  |  |  | **Train** | | **Test** | |
| **Outcome** | **Description** | **Max features (clinical + genetic)** | **Samples** | **Class counts (N)** | **Samples** | **Class counts (N)** |
| OPCRIT 90 | Course of disorder | 107 (86 + 21) | 5,448 | 0 (3,016); 1 (2,432) | 2,331 | 0 (1,291); 1 (1,040) |

GPC: Genomic Psychiatry cohort; AFR: African; AMR: admixed American; EUR: European.

**Supplementary Table 6. Description of train/test splits for Bio*Me* and Genomic Psychiatry Cohort (GPC) datasets.**

| **Bio*Me*** | | | | | | | | | | | | |
| --- | --- | --- | --- | --- | --- | --- | --- | --- | --- | --- | --- | --- |
|  | **AFR** | | | | **AMR** | | | | **EUR** | | | |
|  | **Train** | | **Test** | | **Train** | | **Test** | | **Train** | | **Test** | |
| **Outcome** | **Samples** | **Class counts (N)** | **Samples** | **Class counts (N)** | **Samples** | **Class counts (N)** | **Samples** | **Class counts (N)** | **Samples** | **Class counts (N)** | **Samples** | **Class counts (N)** |
| Aggressive | 246 | 0 (220); 1 (26) | 103 | 0 (93); 1 (10) | 222 | 0 (195); 1 (27) | 94 | 0 (83); 1 (11) | 68 | 0 (63); 1 (5) | 29 | 0 (27); 1 (2) |
| Psychiatric admissions | 245 | 0 (177); 1 (68) | 104 | 0 (75); 1 (29) | 223 | 0 (157); 1 (66) | 93 | 0 (66); 1 (27) | 69 | 0 (51); 1 (18) | 28 | 0 (21); 1 (7) |
| **GPC** | | | | | | | | | | | | |
|  | **AFR** | | | | **AMR** | | | | **EUR** | | | |
|  | **Train** | | **Test** | | **Train** | | **Test** | | **Train** | | **Test** | |
| **Outcome** | **Samples** | **Class counts (N)** | **Samples** | **Class counts (N)** | **Samples** | **Class counts (N)** | **Samples** | **Class counts (N)** | **Samples** | **Class counts (N)** | **Samples** | **Class counts (N)** |
| Course of disorder | 2,466 | 0 (1,545); 1 (921) | 1,056 | 0 (662); 1 (394) | 434 | 0 (229); 1 (205) | 184 | 0 (97); 1 (87) | 2548 | 0 (1,242); 1 (1,306) | 1,091 | 0 (532); 1 (559) |

GPC: Genomic Psychiatry cohort; AFR: African; AMR: admixed American; EUR: European.

**Supplementary Table 7. Description of train/test splits for datasets broken down by ancestry.**

| **Dataset** | **Ancestry** | **Outcome** | **Validation F2 score (sd)** | | | | | **p-val** | **Test F2 score** | | | | | **AUPRC estimate on test set (sd)** | | | | | **p-val** |
| --- | --- | --- | --- | --- | --- | --- | --- | --- | --- | --- | --- | --- | --- | --- | --- | --- | --- | --- | --- |
|  |  |  | **Clinical** | **Clinical and genetic** | **Clinical and binarized genetic** | **Genetic** | **Binarized genetic** |  | **Clinical** | **Clinical and genetic** | **Clinical and binarized genetic** | **Genetic** | **Binarized genetic** | **Clinical** | **Clinical and genetic** | **Clinical and binarized genetic** | **Genetic** | **Binarized genetic** |  |
| Bio*Me* | ALL | Aggressive | 0.653 (0.101) | 0.658 (0.100) | 0.654 (0.102) | 0.129 (0.084) | 0.089 (0.069) | <0.001 | 0.441 | 0.441 | 0.396 | 0.000 | 0.000 | 0.713 (0.080) | 0.712 (0.081) | 0.706 (0.081) | 0.098 (0.012) | 0.087 (0.012) | <0.001 |
| Bio*Me* | ALL | Psychiatric admissions | 0.729 (0.062) | 0.729 (0.057) | 0.728 (0.057) | 0.207 (0.056) | 0.203 (0.055) | <0.001 | 0.672 | 0.682 | 0.668 | 0.094 | 0.057 | 0.773 (0.051) | 0.734 (0.056) | 0.734 (0.056) | 0.294 (0.030) | 0.292 (0.029) | <0.001 |
| GPC | ALL | Course of disorder | 0.555 (0.015) | 0.559 (0.015) | 0.560 (0.015) | 0.389 (0.016) | 0.383 (0.018) | <0.001 | 0.560 | 0.572 | 0.578 | 0.413 | 0.378 | 0.650 (0.013) | 0.659 (0.013) | 0.657 (0.013) | 0.529 (0.014) | 0.518 (0.013) | <0.001 |
| Bio*Me* | AFR | Aggressive | 0.619 (0.180) | 0.610 (0.178) | 0.612 (0.178) | 0.200 (0.150) | 0.178 (0.148) | <0.001 | 0.349 | 0.349 | 0.349 | 0.000 | 0.000 | 0.667 (0.128) | 0.666 (0.128) | 0.674 (0.126) | 0.111 (0.029) | 0.138 (0.057) | <0.001 |
| Bio*Me* | AFR | Psychiatric admissions | 0.693 (0.094) | 0.680 (0.092) | 0.677 (0.092) | 0.196 (0.090) | 0.221 (0.097) | <0.001 | 0.683 | 0.674 | 0.669 | 0.125 | 0.084 | 0.856 (0.054) | 0.828 (0.058) | 0.806 (0.062) | 0.349 (0.063) | 0.336 (0.068) | <0.001 |
| GPC | AFR | Course of disorder | 0.401 (0.023) | 0.410 (0.023) | 0.410 (0.024) | 0.077 (0.023) | 0.066 (0.021) | <0.001 | 0.436 | 0.411 | 0.427 | 0.068 | 0.059 | 0.554 (0.022) | 0.554 (0.023) | 0.553 (0.022) | 0.410 (0.016) | 0.415 (0.017) | <0.001 |
| Bio*Me* | AMR | Aggressive | 0.564 (0.167) | 0.561 (0.167) | 0.556 (0.166) | 0.134 (0.115) | 0.112 (0.113) | <0.001 | 0.490 | 0.490 | 0.490 | 0.208 | 0.106 | 0.698 (0.127) | 0.698 (0.126) | 0.674 (0.128) | 0.393 (0.124) | 0.251 (0.078) | <0.001 |
| Bio*Me* | AMR | Psychiatric admissions | 0.746 (0.092) | 0.741 (0.094) | 0.738 (0.096) | 0.350 (0.105) | 0.327 (0.104) | <0.001 | 0.591 | 0.611 | 0.644 | 0.125 | 0.168 | 0.804 (0.072) | 0.665 (0.090) | 0.659 (0.091) | 0.314 (0.048) | 0.275 (0.036) | <0.001 |
| GPC | AMR | Course of disorder | 0.624 (0.048) | 0.632 (0.048) | 0.630 (0.049) | 0.410 (0.053) | 0.407 (0.053) | <0.001 | 0.545 | 0.565 | 0.567 | 0.426 | 0.436 | 0.623 (0.045) | 0.650 (0.045) | 0.655 (0.045) | 0.482 (0.040) | 0.475 (0.038) | <0.001 |
| Bio*Me* | EUR | Aggressive | 0.050 (0.167) | 0.035 (0.138) | 0.031 (0.132) | 0.028 (0.123) | 0.062 (0.178) | <0.05 | 0.500 | 0.560 | 0.560 | 0.000 | 0.000 | 0.313 (0.296) | 0.494 (0.341) | 0.494 (0.341) | 0.057 (0.021) | 0.057 (0.021) | <0.001 |
| Bio*Me* | EUR | Psychiatric admissions | 0.693 (0.157) | 0.682 (0.153) | 0.660 (0.152) | 0.301 (0.191) | 0.314 (0.196) | <0.001 | 0.606 | 0.606 | 0.588 | 0.303 | 0.303 | 0.645 (0.171) | 0.647 (0.167) | 0.607 (0.175) | 0.448 (0.162) | 0.415 (0.151) | <0.001 |
| GPC | EUR | Course of disorder | 0.693 (0.022) | 0.695 (0.022) | 0.695 (0.022) | 0.840 (0.0001) | 0.840 (0.0001) | <0.001 | 0.669 | 0.668 | 0.670 | 0.840 | 0.840 | 0.692 (0.015) | 0.694 (0.015) | 0.694 (0.015) | 0.574 (0.016) | 0.570 (0.016) | <0.001 |

ALL: all ancestries; AFR: African; AMR: admixed American; EUR: European; PC: principal components; PRS: polygenic risk scores; AUPRC: area under precision-recall curve**.**

**Supplementary Table 8.** **Model** **prediction performances and comparisons.** $F_{2}$ validation estimates and generalization scores are reported. Bootstrap estimate of area under the precision-recall curve (AUPRC) is displayed to investigate models' discriminant ability. P-values refer to one-way repeated-measure ANOVAs run for average comparisons between different feature configurations. Results for clinical, clinical and genetic, and clinical and binarized genetic features are the same as those reported in manuscript’s Table 2 and are repeated here to ease comparisons of all model configurations.

| **Validation F2 score pairwise comparisons (p-val)** | | | | | | | | | | | | | | | **Ancestry** |
| --- | --- | --- | --- | --- | --- | --- | --- | --- | --- | --- | --- | --- | --- | --- | --- |
| Aggressive | Clinical and genetic | Clinical and binarized genetic | Genetic | Binarized genetic | Psychiatric admissions | Clinical and genetic | Clinical and binarized genetic | Genetic | Binarized genetic | Course of disorder | Clinical and genetic | Clinical and binarized genetic | Genetic | Binarized genetic |  |
| Clinical | 0.66 | 0.91 | <0.0001 | <0.0001 | Clinical | 0.96 | 0.85 | <0.0001 | <0.0001 | Clinical | <0.01 | <0.0001 | <0.0001 | <0.0001 | ALL |
| Clinical and genetic | - | 0.72 | <0.0001 | <0.0001 | Clinical and genetic | - | 0.89 | <0.0001 | <0.0001 | Clinical and genetic | - | 0.33 | <0.0001 | <0.0001 |  |
| Clinical and binarized genetic | - | - | <0.0001 | <0.0001 | Clinical and binarized genetic | - | - | <0.0001 | <0.0001 | Clinical and binarized genetic | - | - | <0.0001 | <0.0001 |  |
| Genetic | - | - | - | <0.0001 | Genetic | - | - | - | 0.51 | Genetic | - | - | - | <0.0001 |  |
| Clinical | 0.57 | 0.60 | <0.0001 | <0.0001 | Clinical | 0.12 | <0.05 | <0.0001 | <0.0001 | Clinical | <0.0001 | <0.0001 | <0.0001 | <0.0001 | AFR |
| Clinical and genetic | - | 0.97 | <0.0001 | <0.0001 | Clinical and genetic | - | 0.67 | <0.0001 | <0.0001 | Clinical and genetic | - | 0.96 | <0.0001 | <0.0001 |  |
| Clinical and binarized genetic | - | - | <0.0001 | <0.0001 | Clinical and binarized genetic | - | - | <0.0001 | <0.0001 | Clinical and binarized genetic | - | - | <0.0001 | <0.0001 |  |
| Genetic | - | - | - | 0.08 | Genetic | - | - | - | <0.01 | Genetic | - | - | - | <0.0001 |  |
| Clinical | 0.88 | 0.65 | <0.0001 | <0.0001 | Clinical | 0.63 | 0.39 | <0.0001 | <0.0001 | Clinical | <0.05 | 0.09 | <0.0001 | <0.0001 | AMR |
| Clinical and genetic | - | 0.76 | <0.0001 | <0.0001 | Clinical and genetic | - | 0.69 | <0.0001 | <0.0001 | Clinical and genetic | - | 0.56 | <0.0001 | <0.0001 |  |
| Clinical and binarized genetic | - | - | <0.0001 | <0.0001 | Clinical and binarized genetic | - | - | <0.0001 | <0.0001 | Clinical and binarized genetic | - | - | <0.0001 | <0.0001 |  |
| Genetic | - | - | - | <0.01 | Genetic | - | - | - | <0.01 | Genetic | - | - | - | 0.65 |  |
| Clinical | 0.29 | 0.16 | 0.10 | 0.25 | Clinical | 0.35 | <0.05 | <0.0001 | <0.0001 | Clinical | 0.46 | 0.49 | <0.0001 | <0.0001 | EUR |
| Clinical and genetic | - | 0.74 | 0.57 | <0.05 | Clinical and genetic | - | 0.13 | <0.0001 | <0.0001 | Clinical and genetic | - | 0.97 | <0.0001 | <0.0001 |  |
| Clinical and binarized genetic | - | - | 0.81 | <0.05 | Clinical and binarized genetic | - | - | <0.0001 | <0.0001 | Clinical and binarized genetic | - | - | <0.0001 | <0.0001 |  |
| Genetic | - | - | - | <0.01 | Genetic | - | - | - | 0.32 | Genetic | - | - | - | 1 |  |
| **AUPRC estimate on test set pairwise comparisons (p-val)** | | | | | | | | | | | | | | | **Ancestry** |
| Aggressive | Clinical and genetic | Clinical and binarized genetic | Genetic | Binarized genetic | Psychiatric admissions | Clinical and genetic | Clinical and binarized genetic | Genetic | Binarized genetic | Course of disorder | Clinical and genetic | Clinical and binarized genetic | Genetic | Binarized genetic |  |
| Clinical | 0.95 | 0.68 | <0.0001 | <0.0001 | Clinical | <0.05 | <0.05 | <0.0001 | <0.0001 | Clinical | 0.59 | 0.65 | <0.0001 | <0.0001 | ALL |
| Clinical and genetic | - | 0.73 | <0.0001 | <0.0001 | Clinical and genetic | - | 0.97 | <0.0001 | <0.0001 | Clinical and genetic | - | 0.94 | <0.0001 | <0.0001 |  |
| Clinical and binarized genetic | - | - | <0.0001 | <0.0001 | Clinical and binarized genetic | - | - | <0.0001 | <0.0001 | Clinical and binarized genetic | - | - | <0.0001 | <0.0001 |  |
| Genetic | - | - | - | 0.54 | Genetic | - | - | - | 0.90 | Genetic | - | - | - | 0.48 |  |
| Clinical | 0.95 | 0.67 | <0.0001 | <0.0001 | Clinical | 0.06 | <0.001 | <0.0001 | <0.0001 | Clinical | 1 | 0.97 | <0.0001 | <0.0001 | AFR |
| Clinical and genetic | - | 0.62 | <0.0001 | <0.0001 | Clinical and genetic | - | 0.16 | <0.0001 | <0.0001 | Clinical and genetic | - | 0.97 | <0.0001 | <0.0001 |  |
| Clinical and binarized genetic | - | - | <0.0001 | <0.0001 | Clinical and binarized genetic | - | - | <0.0001 | <0.0001 | Clinical and binarized genetic | - | - | <0.0001 | <0.0001 |  |
| Genetic | - | - | - | 0.07 | Genetic | - | - | - | 0.40 | Genetic | - | - | - | 0.78 |  |
| Clinical | 0.97 | 0.12 | <0.0001 | <0.0001 | Clinical | <0.0001 | <0.0001 | <0.0001 | <0.0001 | Clinical | 0.08 | <0.0001 | <0.0001 | <0.0001 | AMR |
| Clinical and genetic | - | 0.13 | <0.0001 | <0.0001 | Clinical and genetic | - | 0.71 | <0.0001 | <0.0001 | Clinical and genetic | - | 0.74 | <0.0001 | <0.0001 |  |
| Clinical and binarized genetic | - | - | <0.0001 | <0.0001 | Clinical and binarized genetic | - | - | <0.0001 | <0.0001 | Clinical and binarized genetic | - | - | <0.0001 | <0.0001 |  |
| Genetic | - | - | - | <0.0001 | Genetic | - | - | - | <0.05 | Genetic | - | - | - | 0.68 |  |
| Clinical | <0.0001 | <0.0001 | <0.0001 | <0.0001 | Clinical | 0.89 | <0.05 | <0.0001 | <0.0001 | Clinical | 0.88 | 0.88 | <0.0001 | <0.0001 | EUR |
| Clinical and genetic | - | 1 | <0.0001 | <0.0001 | Clinical and genetic | - | <0.01 | <0.0001 | <0.0001 | Clinical and genetic | - | 1 | <0.0001 | <0.0001 |  |
| Clinical and binarized genetic | - | - | <0.0001 | <0.0001 | Clinical and binarized genetic | - | - | <0.0001 | <0.0001 | Clinical and binarized genetic | - | - | <0.0001 | <0.0001 |  |
| Genetic | - | - | - | 1 | Genetic | - | - | - | <0.05 | Genetic | - | - | - | 0.73 |  |

PCs: Principal components; PRS: Polygenic risk scores; AUPRC: area under precision-recall curve; ALL: all ancestries; AFR: African; AMR: admixed American; EUR: European; All: clinical and genetic features; Clinical: clinical features; Genetic: genetic.

**Supplementary Table 9. Post-hoc comparisons for validation** $F_{2}$ **scores and area under precision-recall curve (AUPRC) bootstrap estimates obtained with different feature configurations.** Table includes p-values for two-sided pairwise t-test comparisons with Benjamini-Hochberg correction.

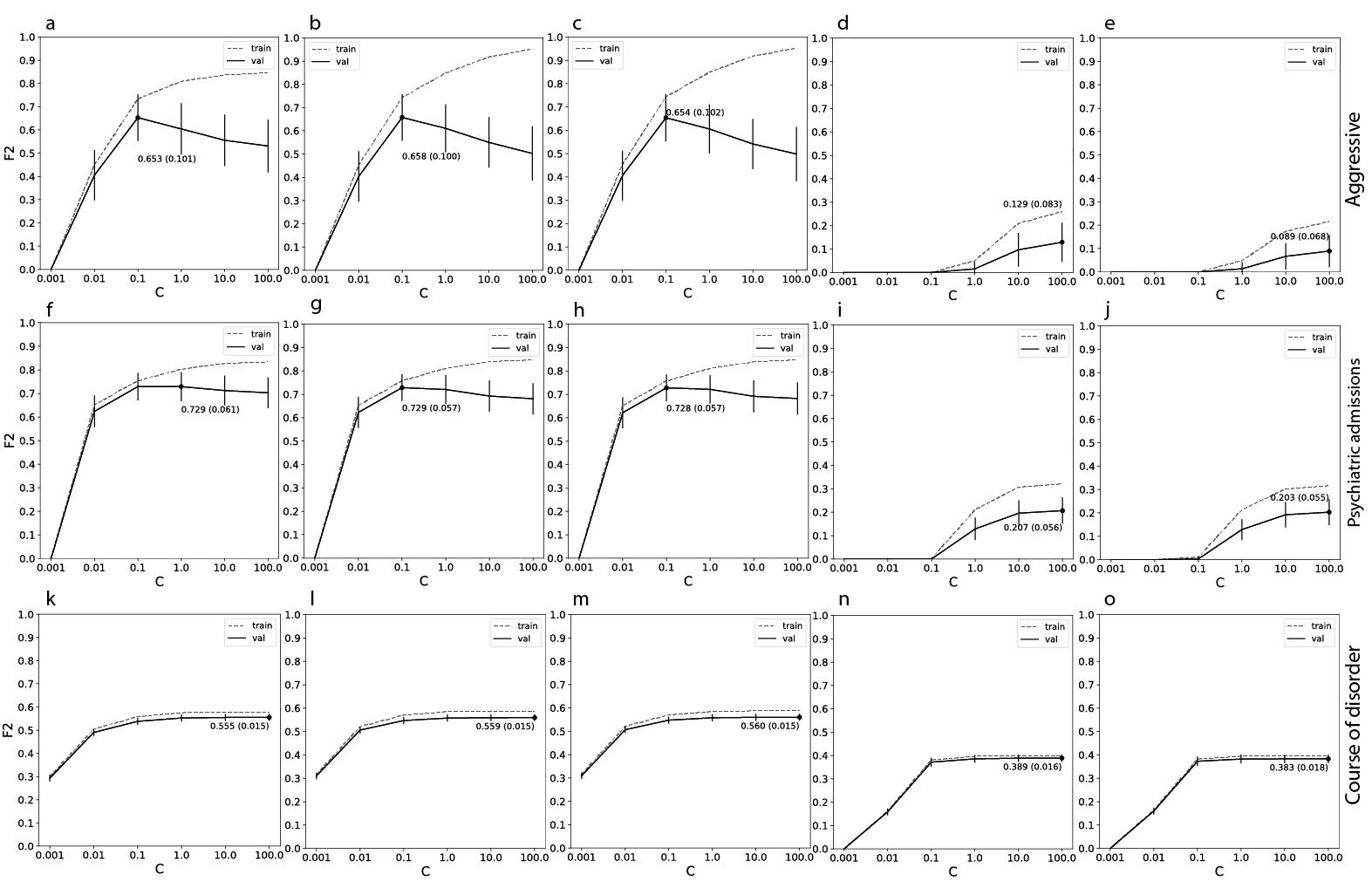

**Supplementary Figure 1. Grid-search for regularization parameter selection within repeated cross-validation framework.** Plots in a row display training and validation $F_{2}$ scores for the prediction of different outcomes with regularization parameter C varying from 0.001 to 100. The dot marks the model with the highest validation score. Plots in column refer to different features included in the model, i.e., clinical (a), (f), (k); clinical and genetic (b), (g), (l); clinical and binarized genetic (c), (h), (m); genetic (d), (i), (n); and binarized genetic (e), (j), (o).

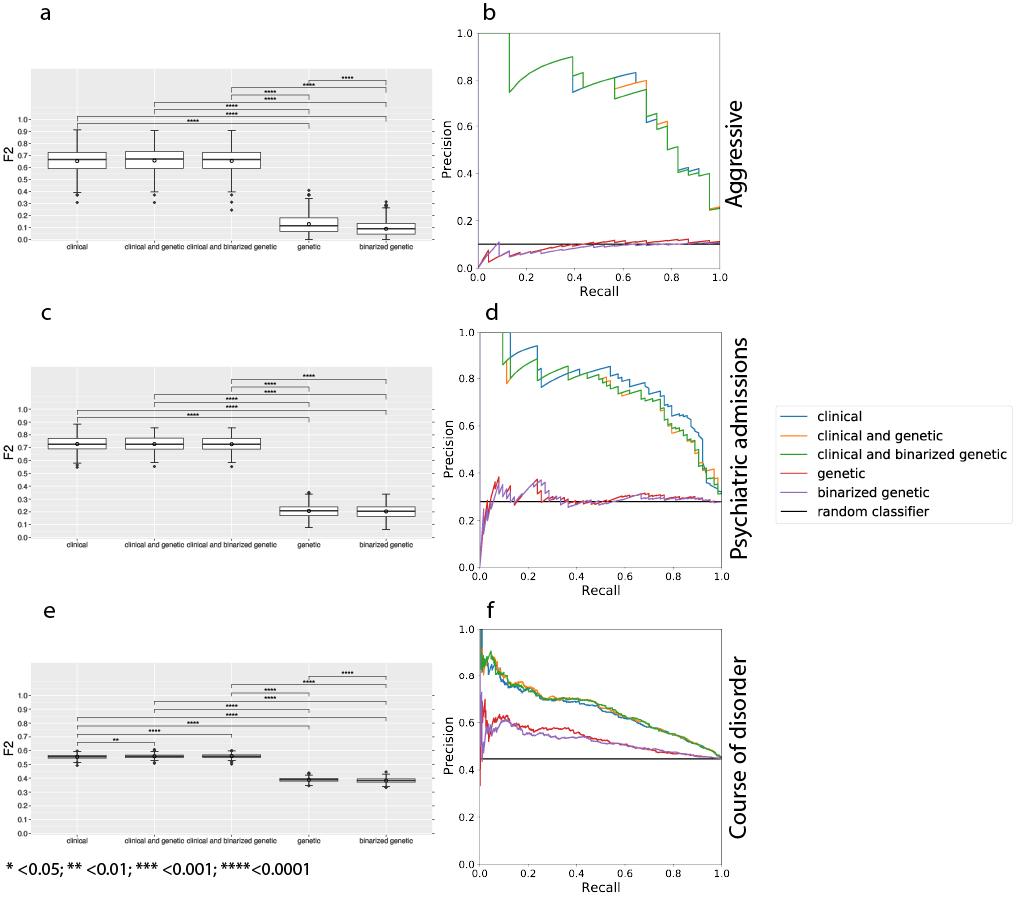

**Supplementary Figure 2. Models’ performance for Bio*Me* and Genomic Psychiatry Cohort (GPC) datasets with different feature configurations.** Average $F_{2}$ scores in predicting different outcomes are displayed in panels (a), (c), and (e). Benjamini-Hochberg corrected p-value thresholds from two-sided pairwise t-test comparisons are displayed for significant results. Validation scores for clinical, clinical and genetic, and clinical and binarized genetic are the same reported in manuscript’s Figure 1 and replicated here to ease comparisons. Panels (b), (d), and (f) display precision-recall curves for linear regression models with different outcomes evaluated on test sets. Performance of random classifiers is displayed as reference.

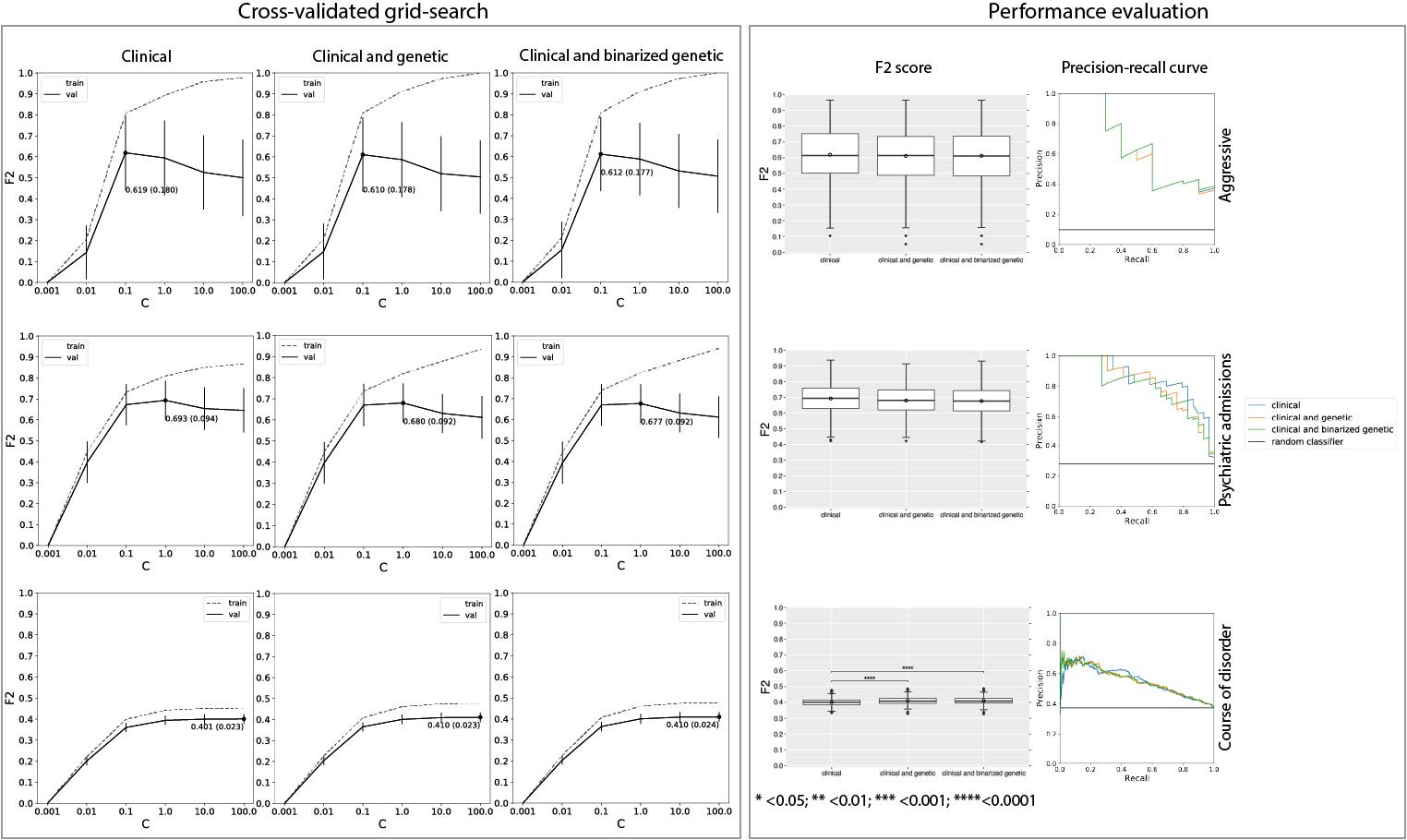

**Supplementary Figure 3. Prediction performance for Bio*Me* and Genomic Psychiatry Cohort (GPC) cohorts restricted to individuals of African (AFR) ancestry.** Training and validation $F_{2}$ estimates for varying regularization parameters (C) are displayed within the “Cross-validated grid-search” frame for each outcome and feature configuration of interest [i.e., clinical, clinical and genetic (all), and clinical and binarized genetic (all binary)]. The dot corresponds to the highest $F_{2}$ score during validation. The best model, with related parameter C, is then trained on the entire training set and evaluated on the test set. Enclosed in the “Performance evaluation” frame are the (1) $F_{2}$ score distributions obtained within repeated cross-validation frameworks. Two-sided pairwise t-test comparison p-values with Benjamini-Hochberg correction are reported (left); and (2) precision-recall curves obtained from the models evaluated on test sets (right).

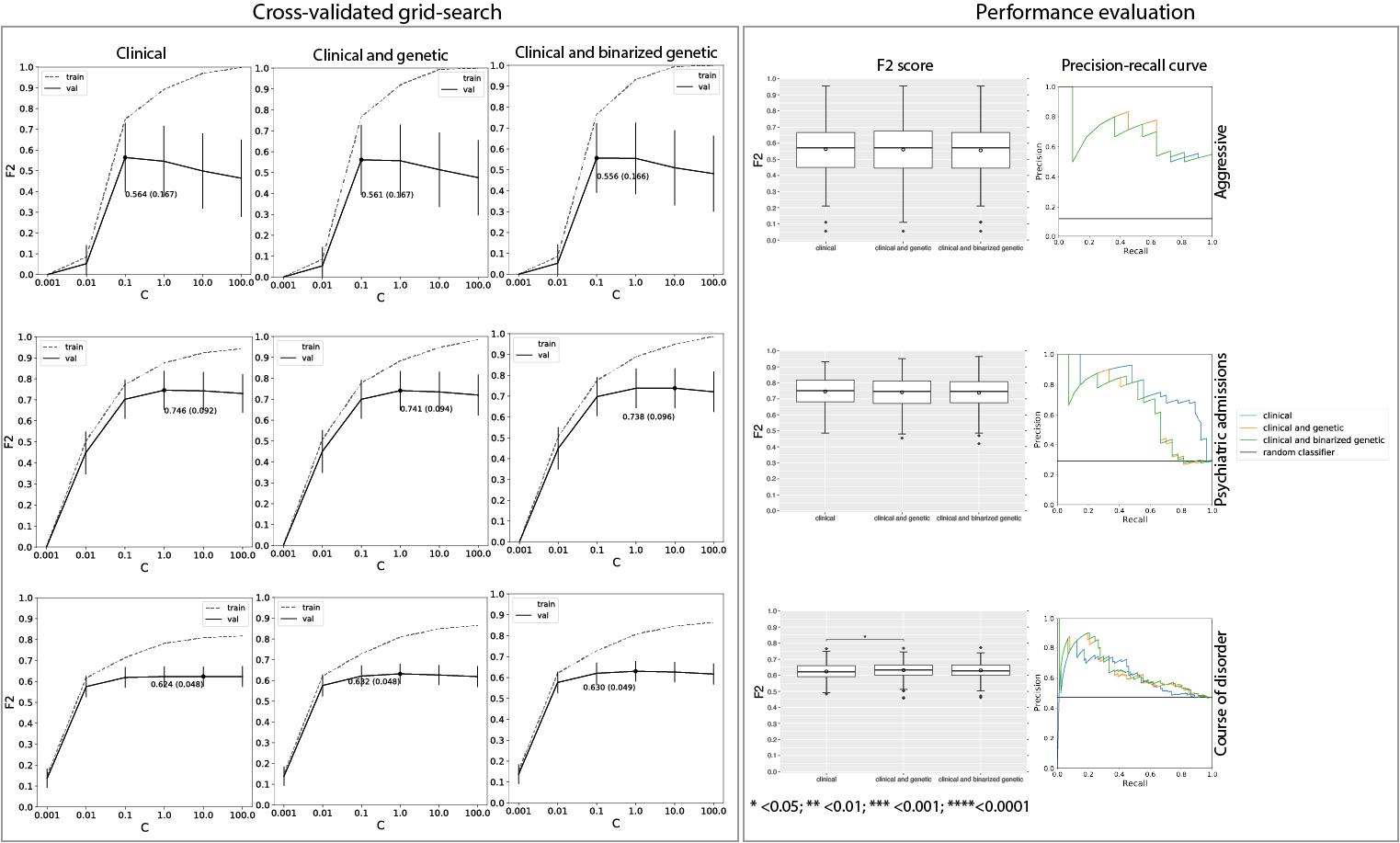

**Supplementary Figure 4. Prediction performance for Bio*Me* and Genomic Psychiatry Cohort (GPC) cohorts restricted to individuals of African (AMR) ancestry.** Training and validation $F_{2}$ estimates for varying regularization parameters (C) are displayed within the “Cross-validated grid-search” frame for each outcome and feature configuration of interest [i.e., clinical, clinical and genetic (all), and clinical and binarized genetic (all binary)]. The dot corresponds to the highest $F_{2}$ score during validation. The best model, with related parameter C, is then trained on the entire training set and evaluated on the test set. Enclosed in the “Performance evaluation” frame are the (1) $F_{2}$ score distributions obtained within repeated cross-validation frameworks. Two-sided pairwise t-test comparison p-values with Benjamini-Hochberg correction are reported (left); and (2) precision-recall curves obtained from the models evaluated on test sets (right).

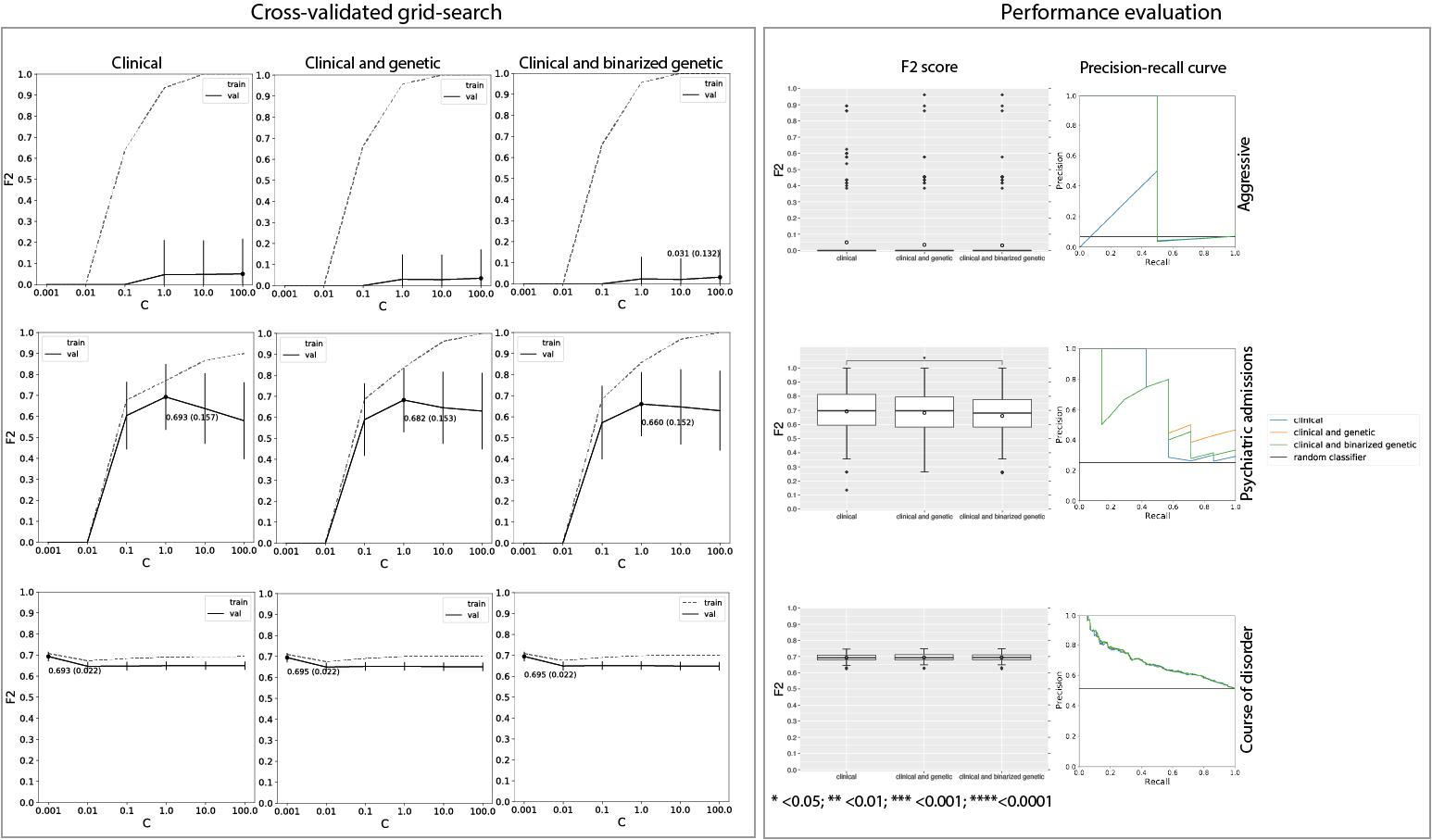

**Supplementary Figure 5. Prediction performance for Bio*Me* and Genomic Psychiatry Cohort (GPC) cohorts restricted to individuals of African (EUR) ancestry.** Training and validation $F_{2}$ estimates for varying regularization parameters (C) are displayed within the “Cross-validated grid-search” frame for each outcome and feature configuration of interest [i.e., clinical, clinical and genetic (all), and clinical and binarized genetic (all binary)]. The dot corresponds to the highest $F_{2}$ score during validation. The best model, with related parameter C, is then trained on the entire training set and evaluated on the test set. Enclosed in the “Performance evaluation” frame are the (1) $F_{2}$ score distributions obtained within repeated cross-validation frameworks. Two-sided pairwise t-test comparison p-values with Benjamini-Hochberg correction are reported (left); and (2) precision-recall curves obtained from the models evaluated on test sets (right).
